# The safety and efficacy of mesenchymal stem cells in the treatment of COVID-19-associated pneumonia: a systematic review and meta-analysis

**DOI:** 10.1101/2021.07.01.21259838

**Authors:** Wang Junwu, Shi Pengzhi, Chen Dong, Wang Shuguang, Wang Pingchuan, Feng Xinmin, Zhang Liang

**Author notes:** **The full contact details of corresponding author Liang Zhang are list below:** Liang Zhang; PhD; Clinical Medical College of Yangzhou University, No.98 Nantong West Road, Yangzhou, 225001, Jiangsu province, China. Co-first authors (Wang Junwu, Shi Pengzhi).

## Abstract

Mesenchymal stem cells (MSCs) therapy is considered one of the most promising treatments in the context of the coronavirus disease 2019 (COVID-19) pandemic. However, the safety and effectiveness of MSCs in the treatment of COVID-19-associated pneumonia patients need to be systematically reviewed and analyzed. Two independent researchers searched for the relevant studies published between October 2019 and April 2021 in PubMed, Embase, Cochrane Library, WAN FANG, and CNKI databases. A total of 22 studies involving 371 patients were included in the present study. MSCs were administered in 247 participants, and MSCs were allogeneic from umbilical cord, adipose tissue, menstrual blood, placenta, Wharton’s jelly, or unreported sources. Combined results found that MSCs group significantly reduced the incidence of adverse events (OR = 0.43, 95%CI. = 0.22∼0.84, P = 0.01) and mortality (OR = 0.17, 95%CI. = 0.06∼0.49, P < 0.01), and the difference compared with control group was statistically significant. No MSCs treat-related serious adverse events were reported. The lung function and radiographic outcomes, and biomarker levels of inflammation and immunity all showed improvement trends. Therefore, MSCs therapy is an effective and safe method in the treatment of COVID-19-associated pneumonia and shows advantages in less adverse events and mortality. However, a standard and effective MSCs treatment program needs to be developed.

## 1 Introduction

Coronavirus disease 2019 (COVID-19), an infectious disease caused by a novel severe acute respiratory syndrome coronavirus 2 (SARS-CoV-2), has been sweeping the globe. According to data reported by the World Health Organization, as of 13 June 2021, 175 333 154 confirmed cases of COVID-19 have been documented in global countries, areas, or territories with 3 793 230 deaths, and 2 655 782 new cases and 72 528 new deaths were reported in the past week [1]. COVID-19 has been associated with an intensive care unit (ICU) admission rate of 5% of proven infections [2] and a high mortality rate of critically ill patients [3]. This series of cruel numbers has prompted an urgent need for treatments that can solve serious cases and prevent fatal consequences [4].

Based on preclinical and clinical studies, mesenchymal stromal cells (MSCs) are found can regulate inflammation and remodeling processes, and restore the concept of alveolar-capillary dysfunction, and thus MSC is considered as a potential treatment for COVID-19 [5,6]. MSCs are pluripotent cells and can be obtained from a variety of tissues, preferably including bone marrow, adipose tissue, placenta, umbilical cord, and dental pulp [7-12]. It is noteworthy that a controlled study by Leng [13] showed that seven patients in MSCs treatment group were cured or the symptoms were significantly improved after 14 days of angiotensin-converting enzyme 2 (ACE2) negative MSCs injection, and inflammation and immune function levels were also ameliorated without observed adverse effects. The outcomes of three patients in control group found one death, one acute respiratory distress syndrome (ARDS), and one remained severely condition. Therefore, they believed that intravenous transplantation of MSCs was safe and effective for the treatment of patients with COVID-19 pneumonia, especially for the critically severe patients. Fortunately, the application of MSCs in the treatment of COVID-19 has been well studied not only in terms of symptomatic efficacy, but also in inflammation, immunity, and molecular mechanisms since then [14,15].

Therefore, this article conducted a systematic review and meta-analysis of the currently available literature on MSCs treatment of COVID-19 since COVID-19 was first reported in 2019, to analyze its safety and efficacy and investigate the potential value of MSCs therapy in patients with COVID-19 infection.

## 2 Methods

### 2.1 Literature Search

The systematic review was performed according to Preferred Reporting Items for Systematic Reviews and Meta-Analyses guidelines [16]. Literature was searched with no language restrictions by two independent researchers. Since COVID-19 was first reported in Wuhan China and subsequently confirmed [17,18], we searched for articles published between October 2019 and April 2021 in PubMed, Embase, Cochrane Library, WAN FANG, and CNKI databases. The terms used for the search were as follows: “novel coronavirus” OR “2019 coronavirus disease” OR “novel coronavirus disease 2019” OR “2019-nCoV” OR “COVID-19” OR “SARS CoV-2” OR “severe acute respiratory syndrome-coronavirus-2” and “stem cell”. Articles from the same authors or institutions were examined, and duplicate data sets were excluded. The number of articles included and excluded was shown in a flow chart (**Fig. 1**).

**Figure 1.**
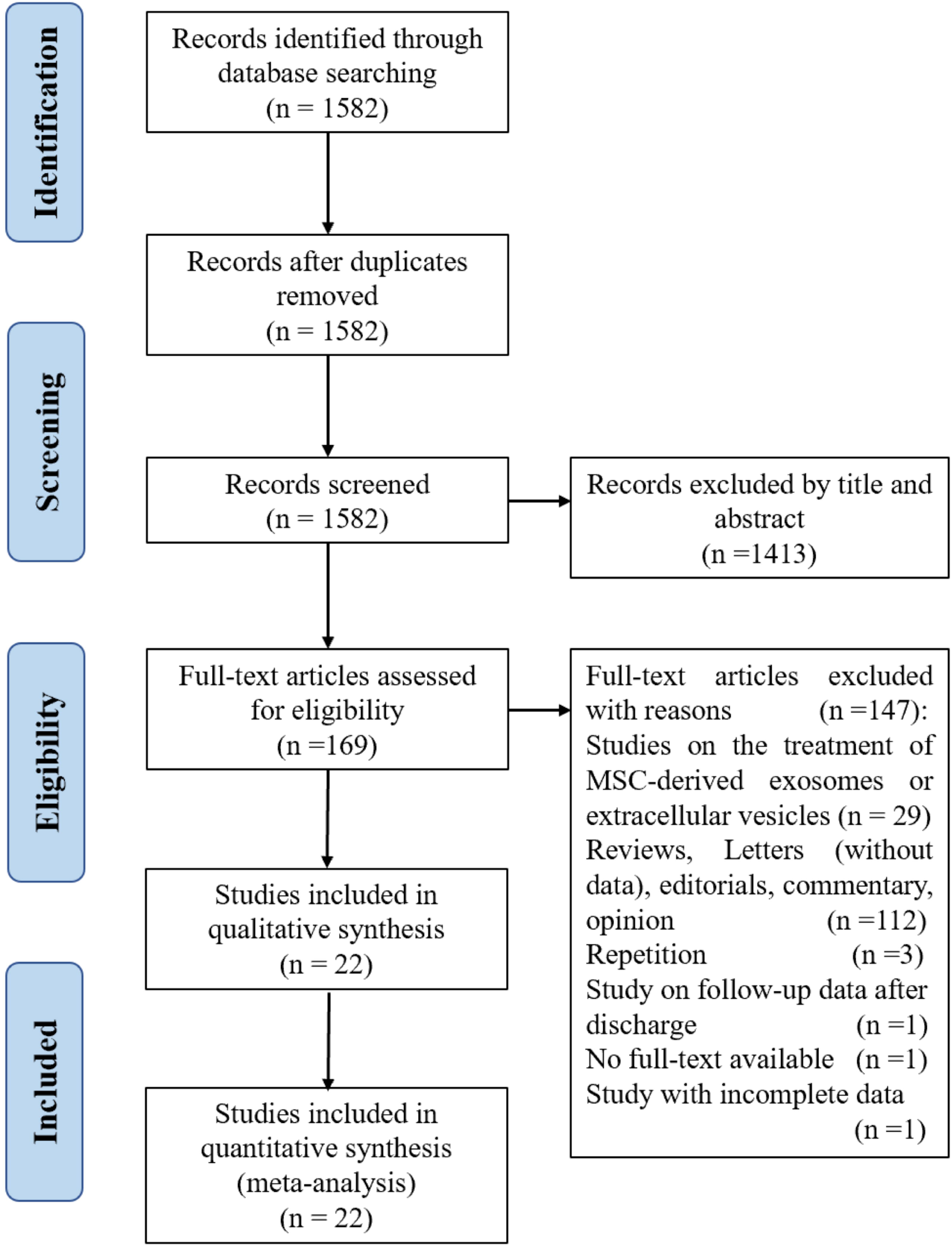
Flow chart

### 2.2 Eligibility Criteria

We included randomized controlled trials (RCT), clinically controlled studies (CCT), retrospective studies, case reports, letters (with valid data), and case series that evaluated the safety and/or efficacy of MSCs administered to adult patients with a diagnosis of COVID-19 pneumonia from any cause. MSCs that were culture-expanded or minimally manipulated were included. Studies were excluded if they did not report original data (eg, reviews, editorials, letters, commentary, opinion, guidelines, or erratum).

### 2.3 Data Extraction

The extracted data were as follows. Data from articles were extracted independently by two reviewers and verified by the third reviewer if there was a disagreement.

#### 2.3.1 General data

The general data were shown in **Table 1-3** (Author name, publication year, country, study design, number of cases, age, gender, and follow-up were shown in **Table 1**; baseline disease severity, comorbidities, general condition, and imaging outcomes were shown in **Table 2**; the MSCs source, surface markers, MSCs dose per time, frequency, cells viability, and transplantation route were shown in **Table 3**). Following established clinical guidelines for the diagnosis and treatment of COVID-19, the disease severity is classified as mild, common, severe, or critical [19,20].

**Table 1.** Study and patient characteristics

| Authors & year<br>& Country | Study design | Total, n<br>MSC; Ctrl | Mean age(year)<br>MSC; Ctrl | Gender, n,<br>(Male/Female)<br>MSC ; Ctrl | Follow-Up |
| --- | --- | --- | --- | --- | --- |
| Leng et al. 2020<br>China [13] | Phase 1,<br>CCT | 10(7; 3) | 57.0; 65.0 | 4/3; 0/3 | 14 days |
| Meng et al. 2020<br>China [22] | Phase 1,<br>CCT | 18(9; 9) | 45.1; 49.6 | 7/2; 4/5 | 28 days |
| Shu et al. 2020 China<br>[23] | RCT | 41(12; 29) | 61.0; 57.9 | 8/4; 16/13 | 28 days |
| Xu et al. 2021 China<br>[24] | Phase 1,<br>CCT | 44(26; 18) | 58.3; 61.1 | 17/9; 13/5 | 30 days |
| Shi et al. 2021 China<br>[25] | Phase 2,<br>RCT | 100(65; 35) | 60.7; 59.9 | 37/28; 19/16 | 28 days |
| Lanzoni et al. 2021<br>United States [26] | Phase 1/2a,<br>RCT | 24(12; 12) | 58.6; 58.8 | 5/7; 8/4 | 31 days |
| Häberle et al. 2021<br>Germany [27] | CCT | 23(5; 18) | 59; 39 | 3/2; 13/5 | NR |
| Sánchez-Guijo et al.<br>2020 Spain [28] | Case series | 13 | 60.3 | 1/12 | 28 days |
| Guo et al. 2020 China<br>[29] | Research<br>Letter | 31 | 70<br>(median) | 25/6 | NR |
| Feng et al. 2020<br>China [30] | Case series | 16 | 61.8 | 12/4 | 28 days |
| Chen et al. 2020<br>China [31] | Retrospective<br>study, Letter | 25 | 70<br>(median) | 20/5 | NR |
| Hashemian et al.<br>2021 Iran [10] | Case series | 11 | 53.8 | 8/3 | 60 days |
| Iglesias et al. 2021<br>Mexico [32] | Case series | 5 | 52.6 | 4/1 | 21 days |
| Yilmaz et al. 2020<br>Turkey [33] | Case report | 1 | 51 | 1/0 | 14 days |
| Zengin et al. 2020<br>Turkey [34] | Case report | 1 | 72 | 1/0 | 60 days |
| Tang et al. 2020<br>China [35] | Case report | 2 | Case 1: 37<br>Case 2: 71 | 1/1 | 14 days |
| Zhang et al.<br>2020 China [12] | Case report | 1 | 54 | 1/0 | 7 days |
| Peng et al. 2020<br>China[36] | Case report | 1 | 66 | 0/1 | 11 days |
| Soler et al. 2020<br>Spain [37] | Letter | 1 | NR | NR | 7 days |
| Zhu et al. 2020 China<br>[38] | Case report | 1 | 48 | 1/0 | 14 days |
| Liang et al. 2020<br>China [39] | Case report | 1 | 65 | 0/1 | 18 days |
| Tao et al. 2020 China<br>[40] | Case report | 1 | 72 | 1/0 | 41 days |

Abbreviations: CCT: clinical controlled trial, **Ctrl**: control group, **MSC**: mesenchymal stem cell treatment group, **RCT**:
randomized controlled trial, NR: not reported

**Table 2.**
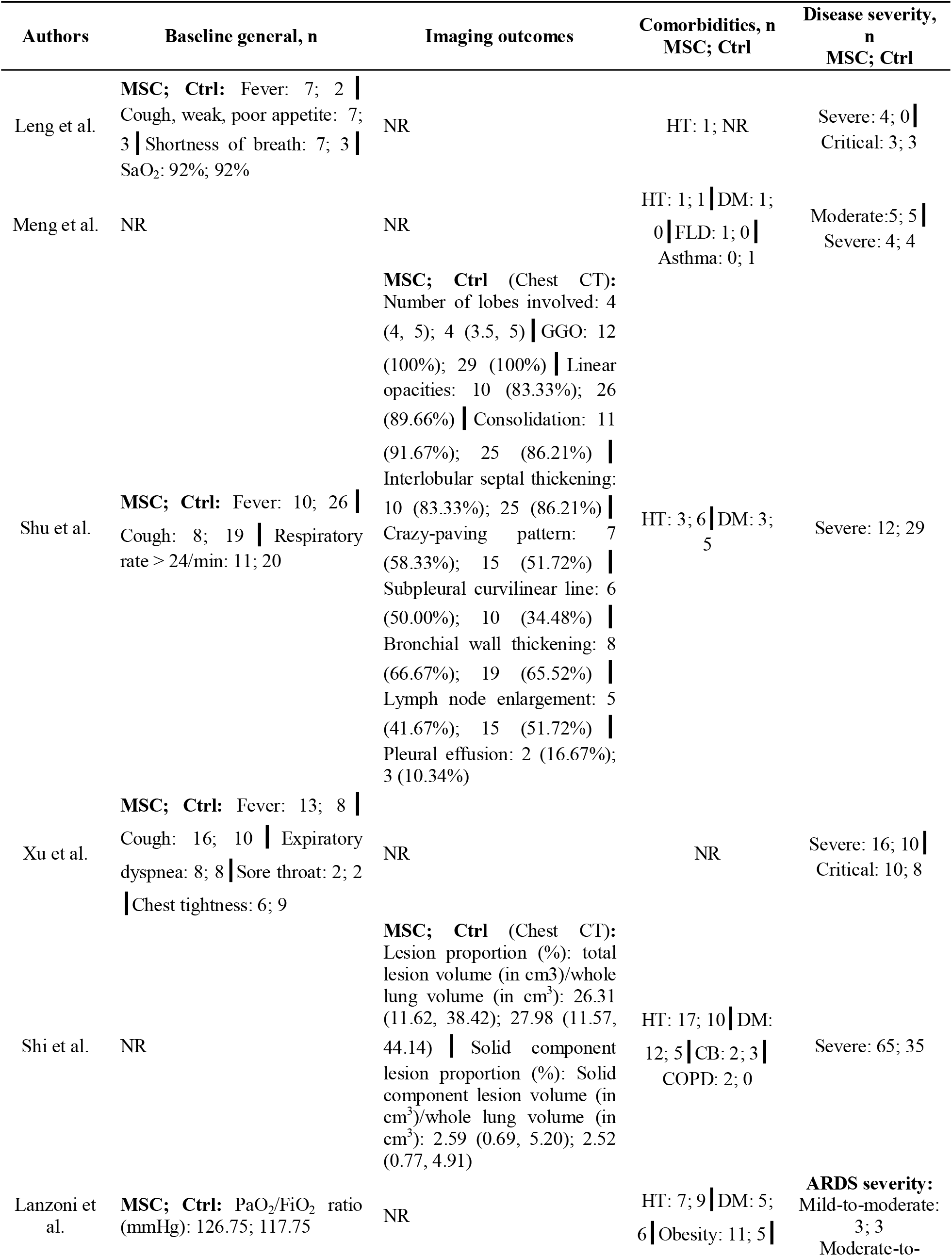
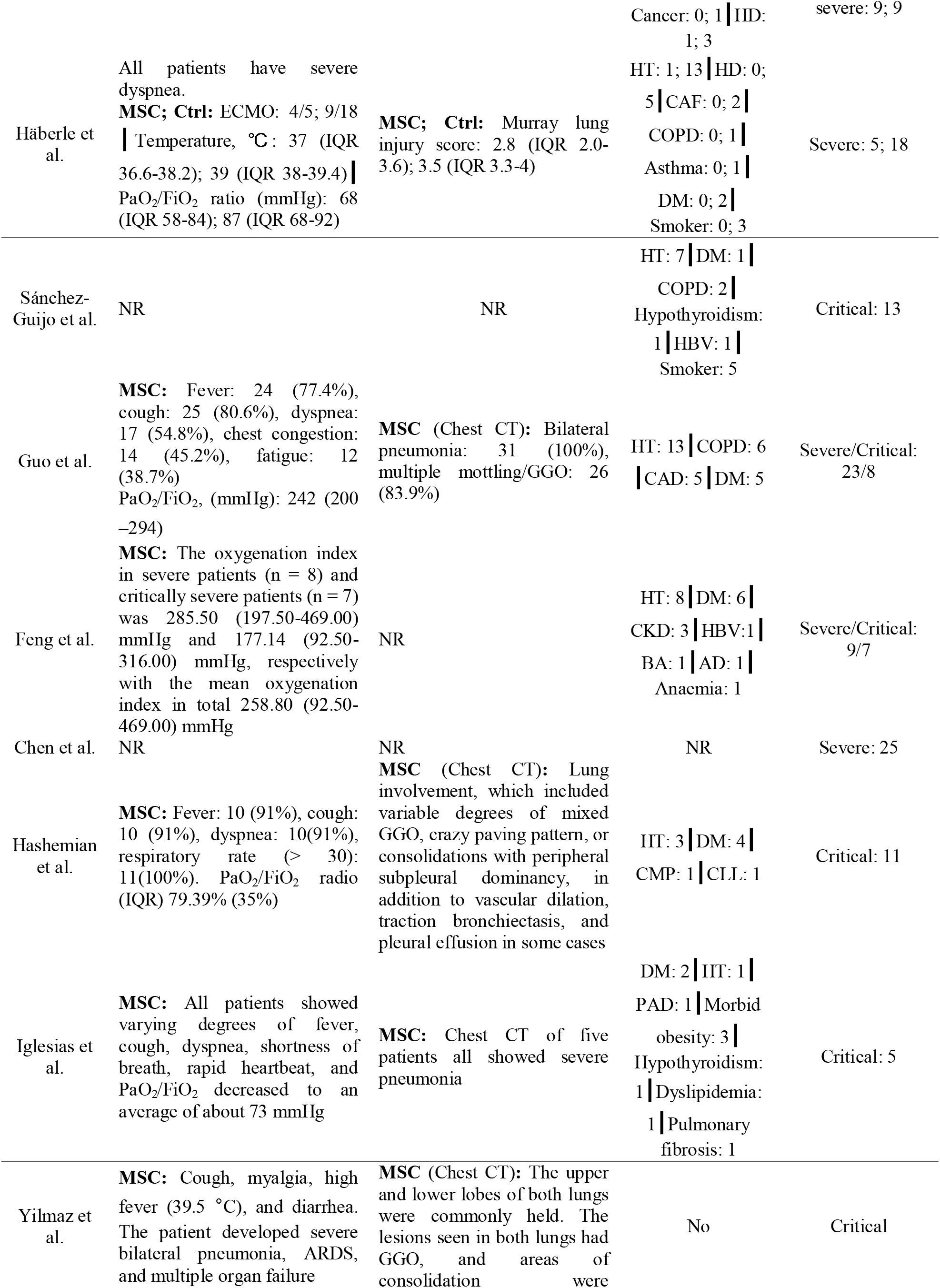
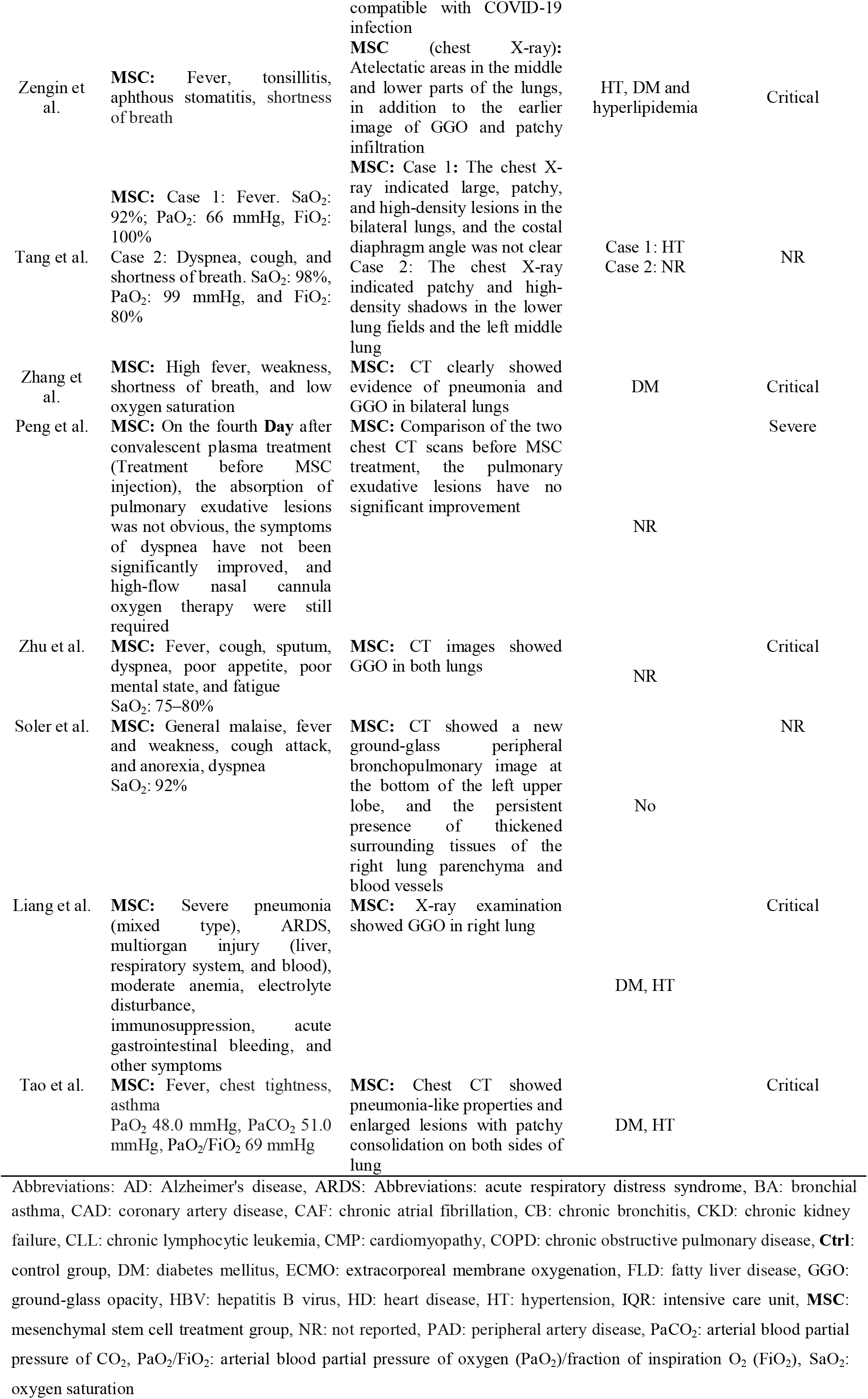
Baseline general, imaging outcomes and disease severity

**Table 3.**
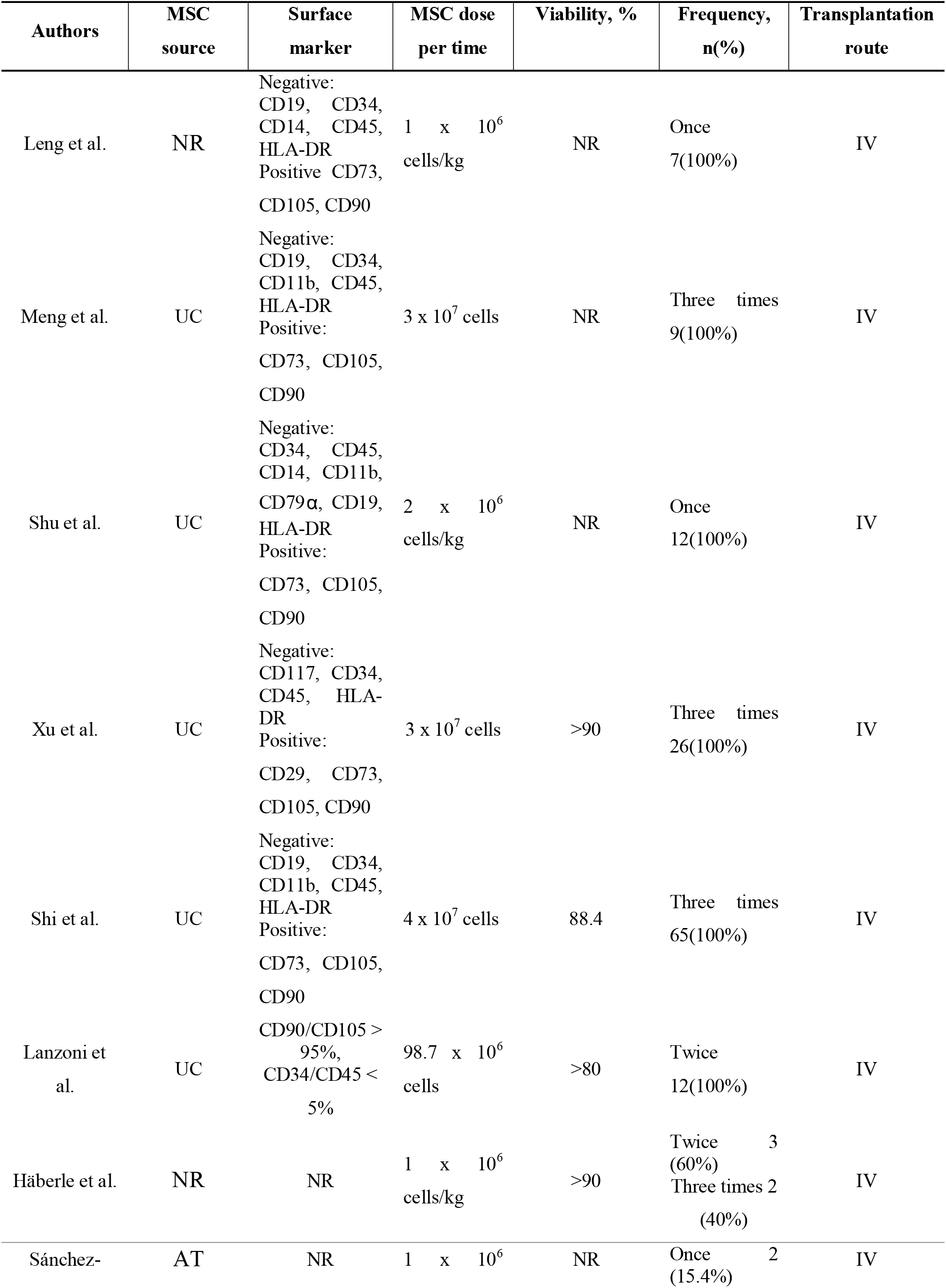
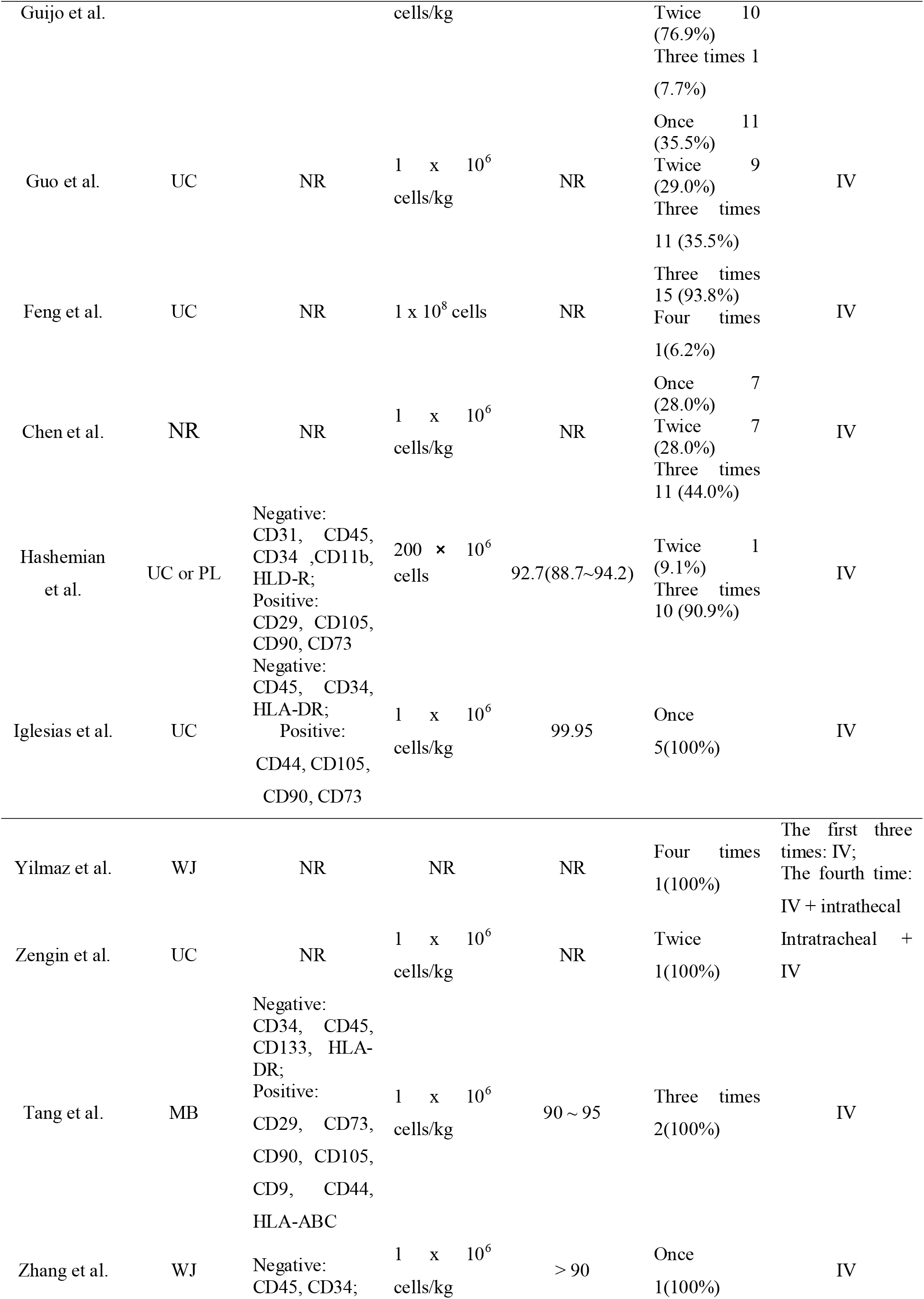
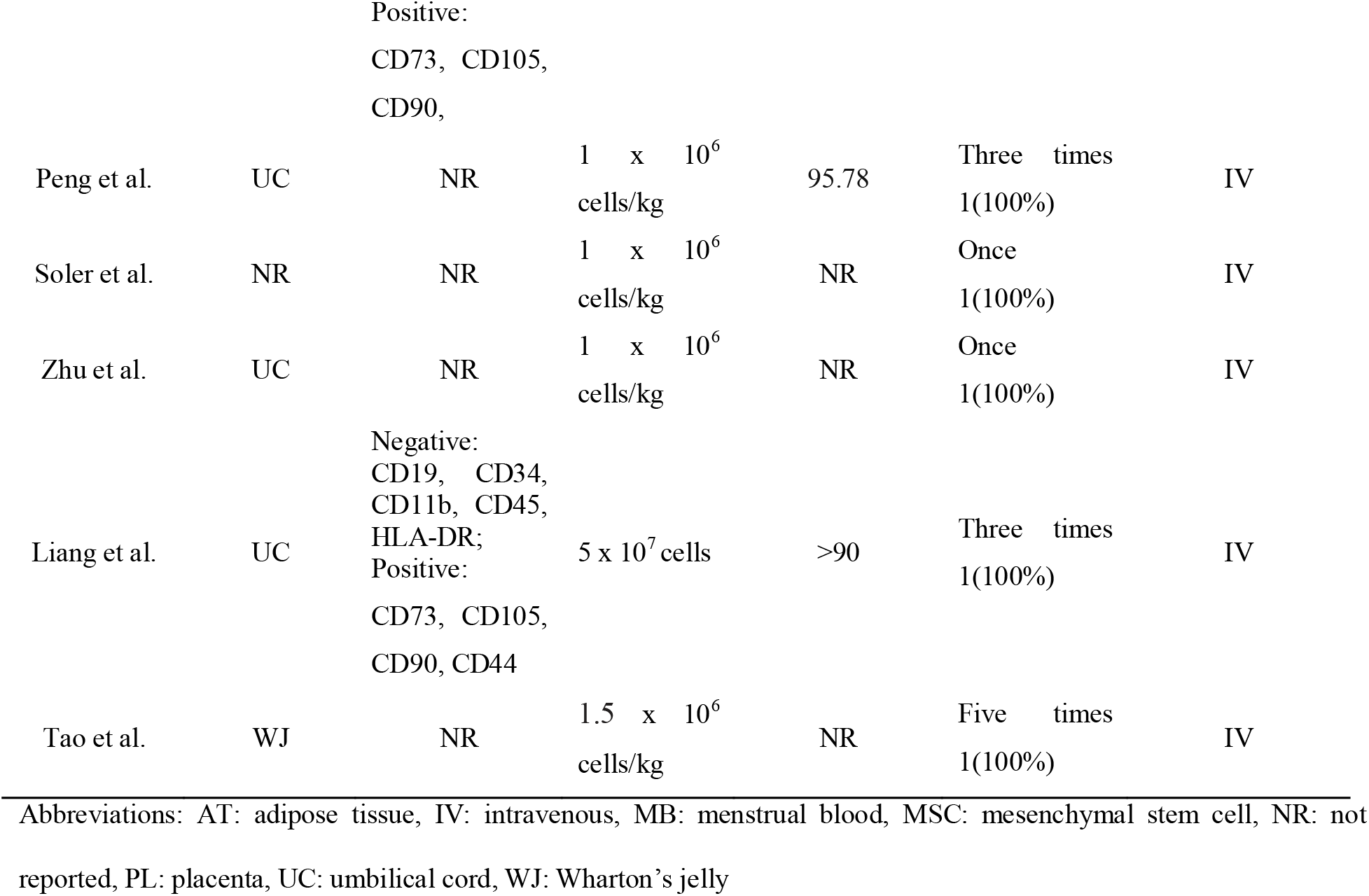
Characteristics of MSCs and intervention methods

#### 2.3.2 Outcomes

The primary outcome was safety based on the number of patients with adverse events (AEs), frequency of AEs, serious AEs (SAEs), and if they were related to the treatment with MSCs. Clinical outcomes included the following: general clinical symptoms (such as fever, cough, dyspnea, respiratory rate, etc), blood oxygen index (arterial blood partial pressure of oxygen (PaO_2_), fraction of inspiration O_2_ (FiO_2_), PaO_2_/FiO_2_, arterial blood or peripheral oxygen saturation (SaO_2_ or SpO_2_), etc), six- or seven-category scale, the time from intervention to recovery, and mortality. Radiographic outcomes: analysis of the lung CT scans or chest X-ray imaging. Laboratory outcomes included the time for nucleic acid turned to be negative, immune cells (dendritic cell (DC), lymphocyte (LYM), natural killer cell (NK cell), T cell, B cell, neutrophil (NE), and white blood cell (WBC)), and inflammatory cytokines (ALT, ammonia, AST, bilirubin, blood creatinine, B-type natriuretic peptide (BNP), blood urea nitrogen (BUN), creatine kinase (CK), CK-MB, C-reactive protein (CRP), cardiac troponin T (cTnT), D-dimer, ferritin, fibrinogen, granulocyte-macrophage colony-stimulating factor (GM-CSF), interferon-γ (INF-γ or IFN-γ), IFN-g, interleukin-1RA (IL-1RA), IL-2, IL-4, IL-5, IL-6, IL-7, IL-8, IL-10, IL-18, IL-22, interferon-inducible protein-10 (IP-10), lactate (LAC); lactate dehydrogenase (LDH), monocyte chemotactic protein-1(MCP-1), macrophage inflammatory protein-1α (MIP-1α), myoglobin, procalcitonin (PCT), platelet derived growth factor-BB (PDGF-BB), regulated upon activation normal T cell expressed and secreted factor (RANTES), tumor necrosis factor-α (TNF-α), TNF-β, triglyceride, and vascular endothelial growth factor (VEGF)).

### 2.4 Quality Assessment

The methodological quality of each study included in the present meta-analysis was evaluated by the National Heart Lung and Blood Institute (NHLBI) quality assessment tools (**Table S1**) [21]. All studies were classified as either good, fair, or poor.

### 2.5 Statistical Analysis

Data are presented as n (%) for categorical variables and mean ± standard deviation for continuous variables. Mortality and the number of patients with AEs were the only two outcomes deemed to be appropriate for meta-analysis. The Review Manager v.5.3 software was used to merge in each study and an overall estimate of the effect was shown in the form of forest plot. We used I^2^ indicator to evaluate heterogeneity between studies. Sensitivity analysis by eliminating one of all included studies at a time and subgroup analysis was performed to examine the source of the heterogeneity when heterogeneity existed (I^2^ > 50%). The random-effects model was used if heterogeneity still existed. Otherwise, the fixed-effects model was used (I^2^ < 50%). The final selected model was used to summarize the odds ratio (OR) of the included studies. We were unable to evaluate publication bias due to the small number of available studies.

## 3 Results

The literature search identified 1582 unique citations. Abstract and full-text screening identified 22 studies with 371 patients to be included for the data extraction. All included studies were assessed as good [10,12,22-26,32,35,36,38,39] or fair [13,27-31,33,34,37,40] according to the NHLBI quality assessment tool (**Table S2**).

### 3.1 Study characteristics

Of the 22 studies [10,12,13,22-40,], there were four CCTs, three RCTs, four case series, three Letters, and eight case reports. Seven were comparative studies with control groups. All 22 studies reported mortality and laboratory outcomes (n = 371); 17 studies reported AEs and SAEs (n = 273); 20 studies reported general clinical symptoms and imaging outcomes after MSCs treatment (n = 324). A total of 247 patients received MSCs therapy, while 124 participated as controls (**Table 1**).

### 3.2 Patient characteristics

The 22 studies were from seven countries and regions, including China (n = 14) [12,13,22-25,29-31, 35,36,38-40], the United States (n = 1) [26], Germany (n = 1) [27], Spain (n = 2) [28,37], Iran (n = 1) [10], Mexico (n = 1) [32], and Turkey (n = 2) [33,34]. Eleven studies reported on baseline oxygenation indicators of patients [10,13,26,27,29,30,32,35,37,38,40]. General symptoms such as fever, cough, and dyspnea were reported in 16 studies [10,12,13,23,24,27,29,32-40], and lung imaging evaluation showed COVID-19 related pneumonia in 15 studies [10,12,23,25,27,29,32-40]. The disease severity of the patient’s condition was Critical (n = 63), Severe (n = 260), and Moderate (n = 10) in 19 studies [10,12,13,22-25,27-34,36,38-40]; and the patients in one study were divided into “Mild-to-moderate” (n = 6) and “Moderate-to-severe” (n = 18) according to the severity of ARDS [26]. The average age of study participants was 45.1 to 61.0 years for MSCs group and 39.0 to 65.0 years for control group in the comparative studies[13,22-27]. In the comparative studies, the disease severity of MSCs group were Critical (n = 13), Severe (n = 115), and Moderate (n = 8), and those of control group were Critical (n = 11), Severe (n = 105), and Moderate (n = 8) [13,22-27]. The most common comorbidities including hypertension (HT, n = 105), diabetes mellitus (DM, n = 61), obesity(n = 19), chronic obstructive pulmonary disease (COPD, n = 11), and heart disease (HD, n = 9) had been reported in 18 studies [10,12,13,22,23,25-30,32-35,38-40]. There were also differences in the follow-up time after MSCs treatment in these studies, ranging from a week to two months, which mainly due to the point when patients were recovery or died and discharge from the hospital (**Table 1-2**).

### 3.3 Intervention method

Culture-expanded allogeneic MSCs were used in all 22 included studies. Allogeneic umbilical cord-derived MSCs were used in 13 studies [22-26,29,30,32,34,36-39], Wharton’s jelly-derived MSCs in three studies [12,33,40], menstrual blood- or adipose tissue- or placenta-derived MSCs were separately used in one study [10,28,35]. Four studies did not report the tissue origin of the MSCs [13,27,31,37](**Table 3**). Characterization was reported in most studies, with significant differences in details. Characterization of MSCs was reported in 11 studies, with most of the following markers: positive for CD9, CD29, CD44, CD73, CD90, CD105, HLA-ABC, and negative for CD11b, CD14, CD19, CD31, CD34, CD45, CD79α, CD133 and HLA-DR [10,12,13,22-26,32,35,39]. Viability was reported to be >80% [10,12,24-27,32,35,36,39].

MSCs were mainly injected intravenously (n = 245), and two other patients were injected intratracheally and intravenously [33,34]. MSCs infusion frequency ranged from a single administration to five administrations (n = 47, 43, 154, 2, 1). Dosing of MSCs ranged from 1 to 2 million cells per kilogram of body weight or a uniform dose of 30 to 200 million cells. Only one study did not report the dose [33].

### 3.4 Safety

#### 3.4.1 Adverse events

We extracted data from 16 studies on the number of AEs and the number of patients with AEs to describe the occurrence of AEs (**Table 4**) [10,12,13,22,24-26,28,31-34,36-39]. In MSCs group, patients with AEs accounted for 48.33% (87/180), and the total number of AEs was 167. Additionally, according to research by Feng et al, hypoalbuminemia, insomnia, gastrointestinal diseases, and paroxysmal arrhythmia occurred in the surviving patients [30]. It is worth noting that 15 treatment-related AEs were reported in five studies. Meng et al reported that two patients receiving MSCs developed transient facial flushing and fever immediately on infusion, which resolved spontaneously within 4 h; another moderate patient had a transient fever within 2 h that resolved within 24 h [22]. Chen et al reported that three cases experienced treatment-related AEs, specifically liver dysfunction, heart failure and allergic rash [31]. Hashemian et al reported that two cases developed shivering that occurred during the initial MSCs infusion, which was relieved by supportive treatment in less than 1 h [10]. Iglesias et al reported that a patient had muscle contractions in extremities, and another patient had muscle contractions in extremities and chest, PO_2_ decreased 78%, arterial hypertension, and respiratory effort; the third patient developed hypotension [32]. In the study of Lanzoni et al, a treatment-related AE was reported without specific description [26]. Comparisons of AEs incidence between MSCs group and control group were made in five studies (n = 196) [13,22,24-26,]. The overall AEs incidences were 58.0% (69/119) for MSCs-treated patients and 77.9% (60 of 77) for controls, and the difference was statistically significant (P = 0.01,OR = 0.43, 95%CI. = 0.22∼0.84) (**Fig. 2**).

**Table 4.**
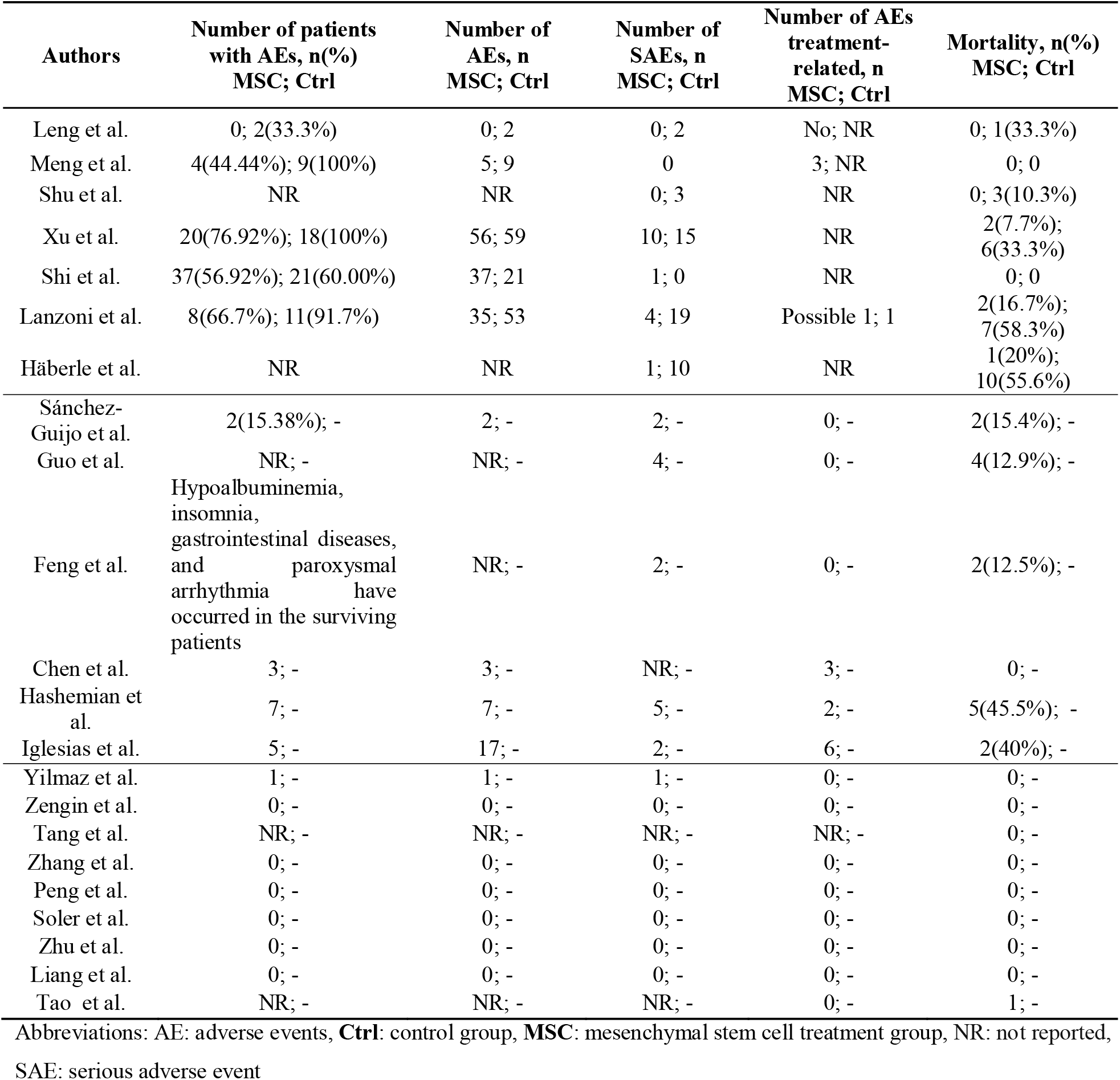
Adverse events and mortality

| Authors | Number of patients<br>with AEs, n(%)<br>MSC; Ctrl | Number of<br>AEs, n<br>MSC; Ctrl | Number of<br>SAEs, n<br>MSC; Ctrl | Number of AEs<br>treatment-related, n<br>MSC; Ctrl | Mortality, n(%)<br>MSC; Ctrl |
| --- | --- | --- | --- | --- | --- |
| Leng et al. | 0; 2(33.3%) | 0; 2 | 0; 2 | No; NR | 0; 1(33.3%) |
| Meng et al. | 4(44.44%); 9(100%) | 5; 9 | 0 | 3; NR | 0; 0 |
| Shu et al. | NR | NR | 0; 3 | NR | 0; 3(10.3%) |
| Xu et al. | 20(76.92%); 18(100%) | 56; 59 | 10; 15 | NR | 2(7.7%);<br>6(33.3%) |
| Shi et al. | 37(56.92%); 21(60.00%) | 37; 21 | 1; 0 | NR | 0; 0 |
| Lanzoni et al. | 8(66.7%); 11(91.7%) | 35; 53 | 4; 19 | Possible 1; 1 | 2(16.7%);<br>7(58.3%) |
| Häberle et al. | NR | NR | 1; 10 | NR | 1(20%);<br>10(55.6%) |
| Sánchez-Guijo et al. | 2(15.38%); - | 2; - | 2; - | 0; - | 2(15.4%); - |
| Guo et al. | NR; - | NR; - | 4; - | 0; - | 4(12.9%); - |
| Feng et al. | Hypoalbuminemia,<br>insomnia,<br>gastrointestinal diseases,<br>and paroxysmal<br>arrhythmia have<br>occurred in the surviving<br>patients | NR; - | 2; - | 0; - | 2(12.5%); - |
| Chen et al. | 3; - | 3; - | NR; - | 3; - | 0; - |
| Hashemian et<br>al. | 7; - | 7; - | 5; - | 2; - | 5(45.5%); - |
| Iglesias et al. | 5; - | 17; - | 2; - | 6; - | 2(40%); - |
| Yilmaz et al. | 1; - | 1; - | 1; - | 0; - | 0; - |
| Zengin et al. | 0; - | 0; - | 0; - | 0; - | 0; - |
| Tang et al. | NR; - | NR; - | NR; - | NR; - | 0; - |
| Zhang et al. | 0; - | 0; - | 0; - | 0; - | 0; - |
| Peng et al. | 0; - | 0; - | 0; - | 0; - | 0; - |
| Soler et al. | 0; - | 0; - | 0; - | 0; - | 0; - |
| Zhu et al. | 0; - | 0; - | 0; - | 0; - | 0; - |
| Liang et al. | 0; - | 0; - | 0; - | 0; - | 0; - |
| Tao et al. | NR; - | NR; - | NR; - | 0; - | 1; - |

Abbreviations: AE: adverse events, **Ctrl**: control group, **MSC**: mesenchymal stem cell treatment group, NR: not reported,
SAE: serious adverse event

**Figure 2.**
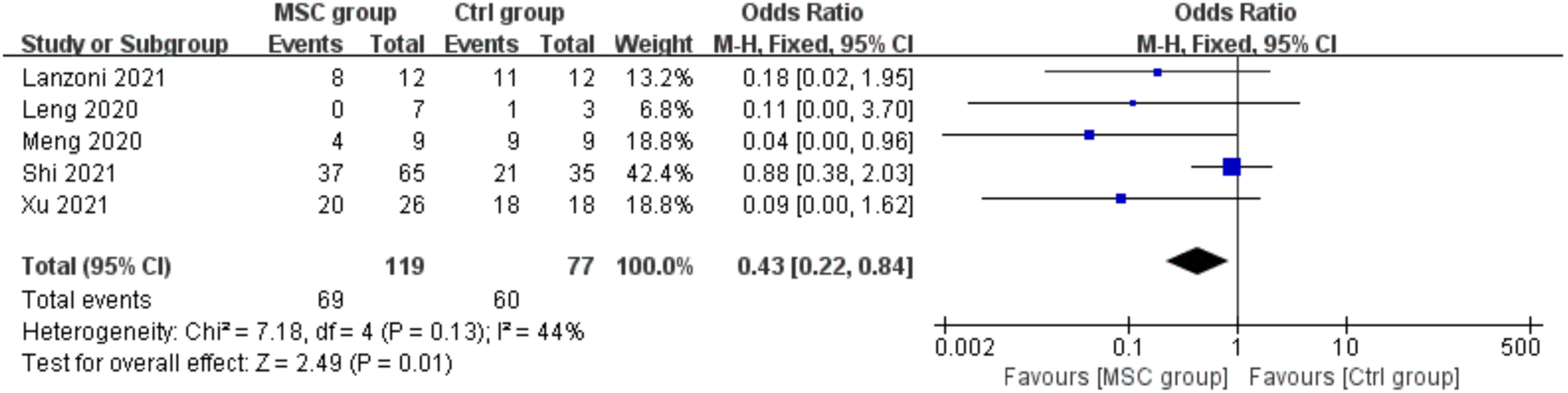
Pooled estimate for the number of adverse events

#### 3.4.2 Serious adverse events

In our data extraction process, the number of deaths was included in the number of SAEs if the study did not describe SAEs in detail and only reported the mortality. The results showed that 32 SAEs occurred in the 219 MSCs-treated patients in 19 studies (**Table 4**) [10,12,13,22-30,32-34,36-39]. There were no reported MSCs treatment-related SAEs. The 10 SAEs reported in Xu’s study included severe liver dysfunction (n = 1), expiratory dyspnea (n = 1), respiratory failure (n = 1), ARDS (n = 1), shock (n = 3), multifunctional organ failure (n = 2), and gastrointestinal bleeding (n = 1) [24]. Shi et al reported 1 case of pneumothorax in MSCs group, and the patient recovered naturally after conservative treatment [25]. Lanzoni et al reported 4 SAEs without detailed introduction [26]. Häberle et al reported a death case due to multiple organ failure [27]. Sanchez-Guijo et al reported that two patients died, one from massive gastrointestinal bleeding and another one from secondary fungal pneumonia by Saccharomyces spp [28]. Guo et al reported 4 patients died without detailed introduction [29]. There were two SAEs during the trial reported by Feng et al [30]. The two patients suffered from bacterial pneumonia and septic shock and died of multiple organ failure or circulation and respiratory failure, respectively. Hashemian et al reported deaths of 4 patients due to multiple organ failure and 1 patient due to cardiac arrest [10]. Iglesias et al reported that a case developed to left lower extremity arterial thrombosis, deterioration of hemodynamics, D-dimer concentration of 7268 ng/mL and death; Enterobacter cloacae were cultured in aspirated samples from his trachea. The other patient developed hemodynamic alterations, epistaxis, hematuria, and died 13 days after MSCs infusion [32]. In the case report of Yilmaz, a patient was diagnosed with upper gastrointestinal bleeding, but his vital signs were stable after effective treatment [33]. Among the seven controlled studies, 136 patients in MSC groups had 16 SAEs, while 124 patients in control group had 49 SAEs [13,22-27].

### 3.5 Efficacy

#### 3.5.1 Mortality

Mortality was reported in all included studies (**Table 4**), and the overall mortality rate was 12.94% (48/371). The mortality rate among MSCs-treated patients was 8.50% (21/247). Comparisons between MSCs group and control group were made in seven studies (n = 260), and there was a trend toward a decreased mortality rate in MSCs group in all seven studies [13,22-27]. The overall mortality rate was 3.68% (5/137) for MSCs-treated patients and 21.77% (27/124) for controls. There was a favorable trend and the difference was statistically significant (P < 0.01, OR = 0.17, 95%CI. = 0.06∼0.49). However, the certainty of the evidence on the impact on mortality was limited due to the differences in the baseline conditions of the included patients, and the imprecision and methodological limitations of the included trials (**Fig. 3, Table 1-2**).

**Figure 3.**
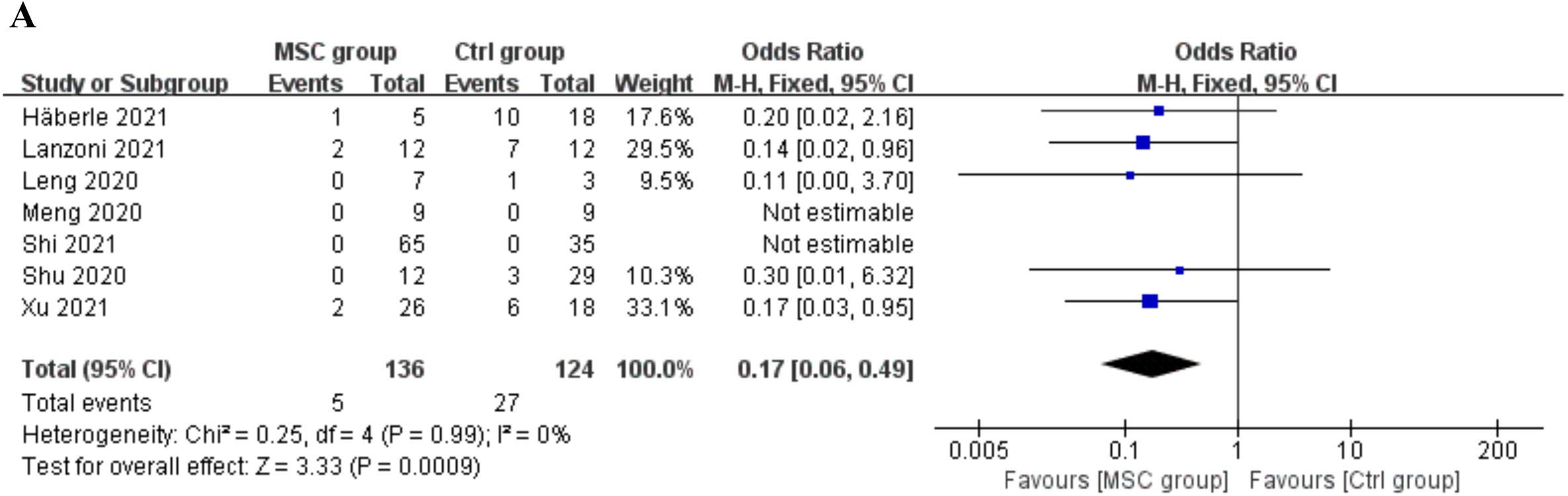
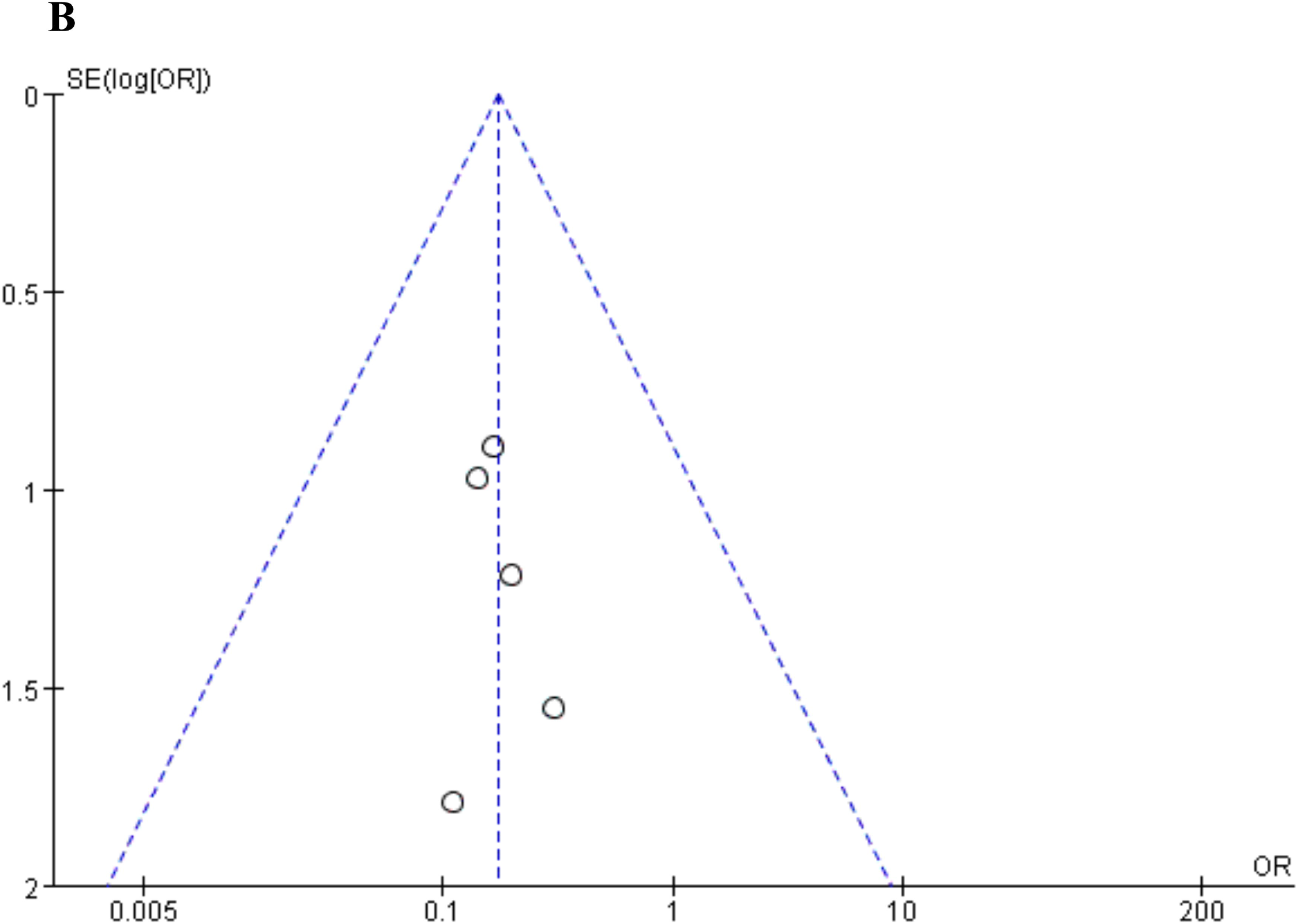
Pooled estimate for mortality. **A** Forest plots of mortality. **B** Funnel plot of mortality

#### 3.5.2 Changes in general clinical symptoms and lung function

The results found that the average time from receiving the first injection to recovery or discharge from hospital ranged from about 2 to 24 days for MSCs-treated patients (**Table 5**) [10,12,13,22-24,26,28,32, 36-40]. In several controlled studies, Shu et al, Xu et al and Lanzoni et al reported that the average time taken to improve (or recovery) for MSCs group was shorter than that of control group, and the difference was statistically significant [23,24,26]. However, Meng et al reported that the duration from admission to discharge between the two groups was close [22]; and Xu et al reported that there was no significant difference in either the length of hospital stay or the number of days in the ICU between the two groups [24].

**Table 5.**
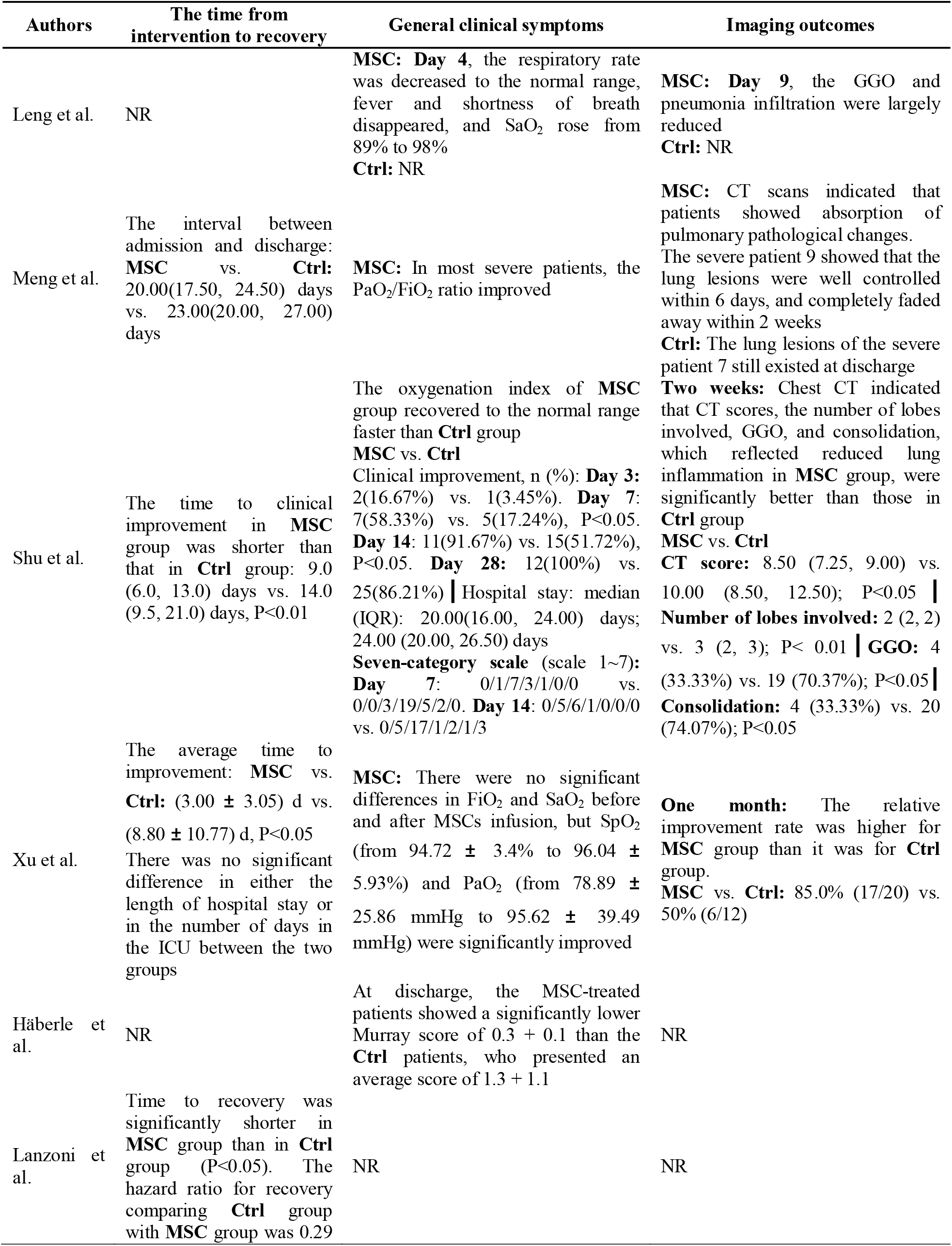
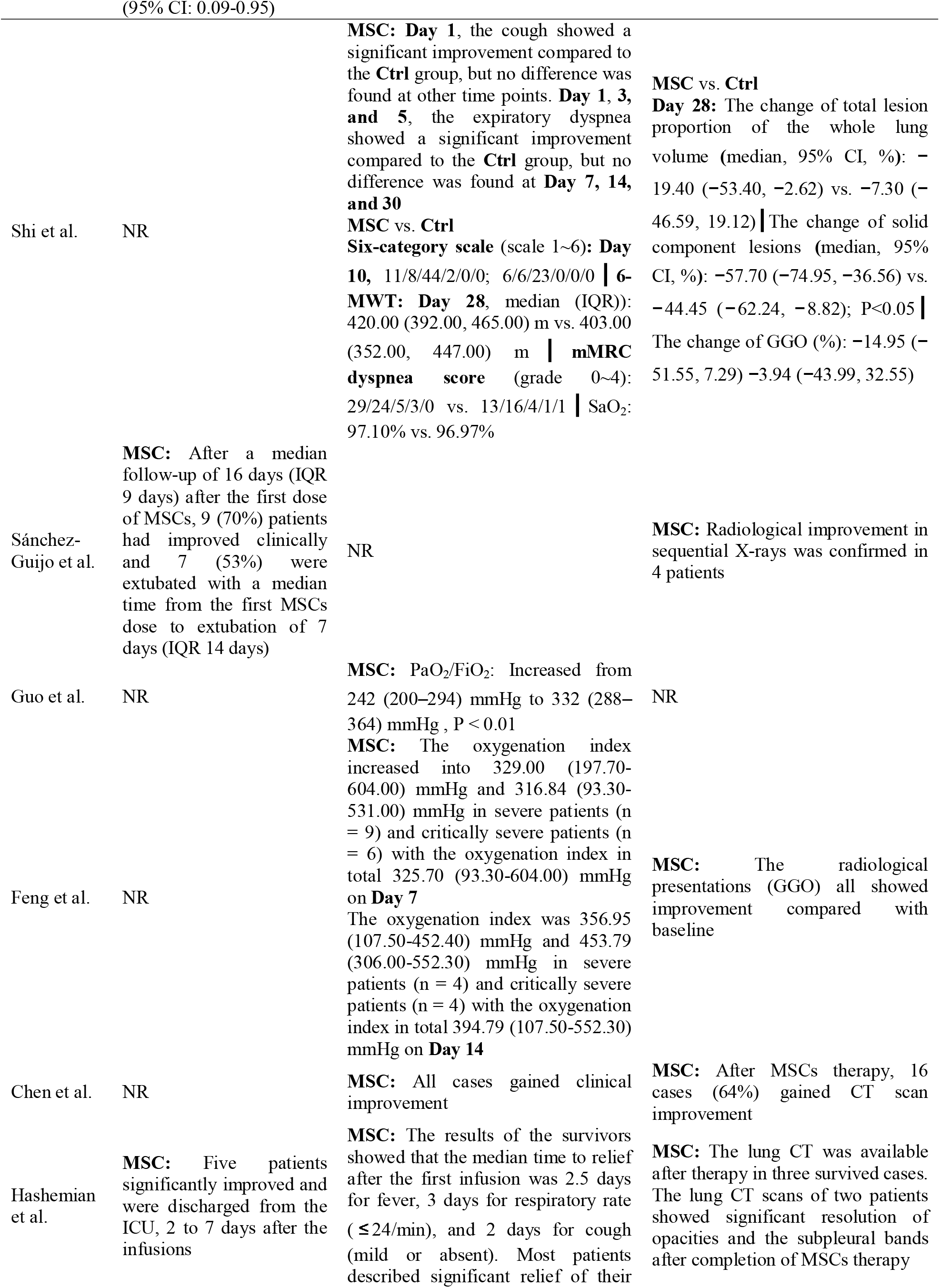
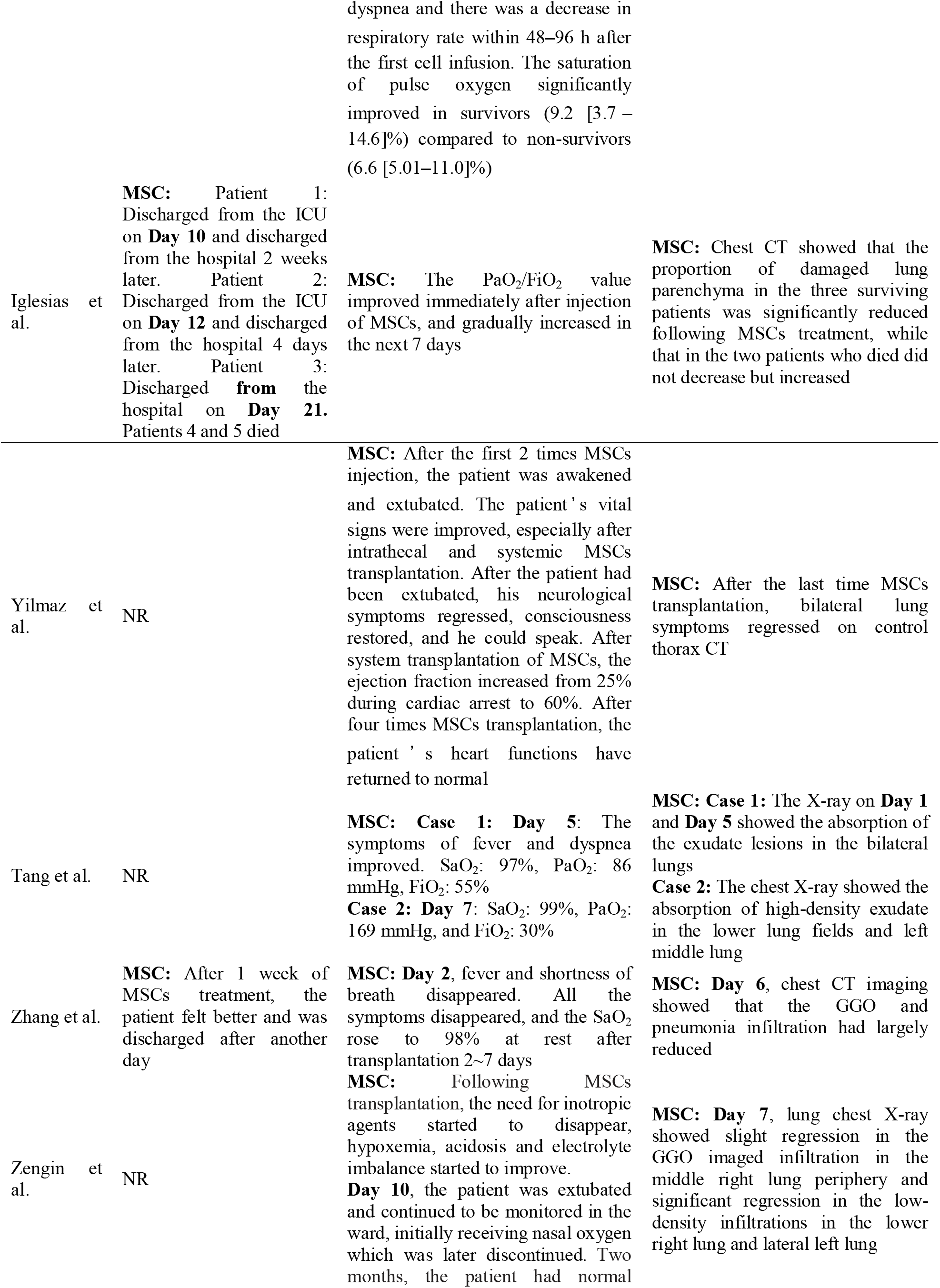
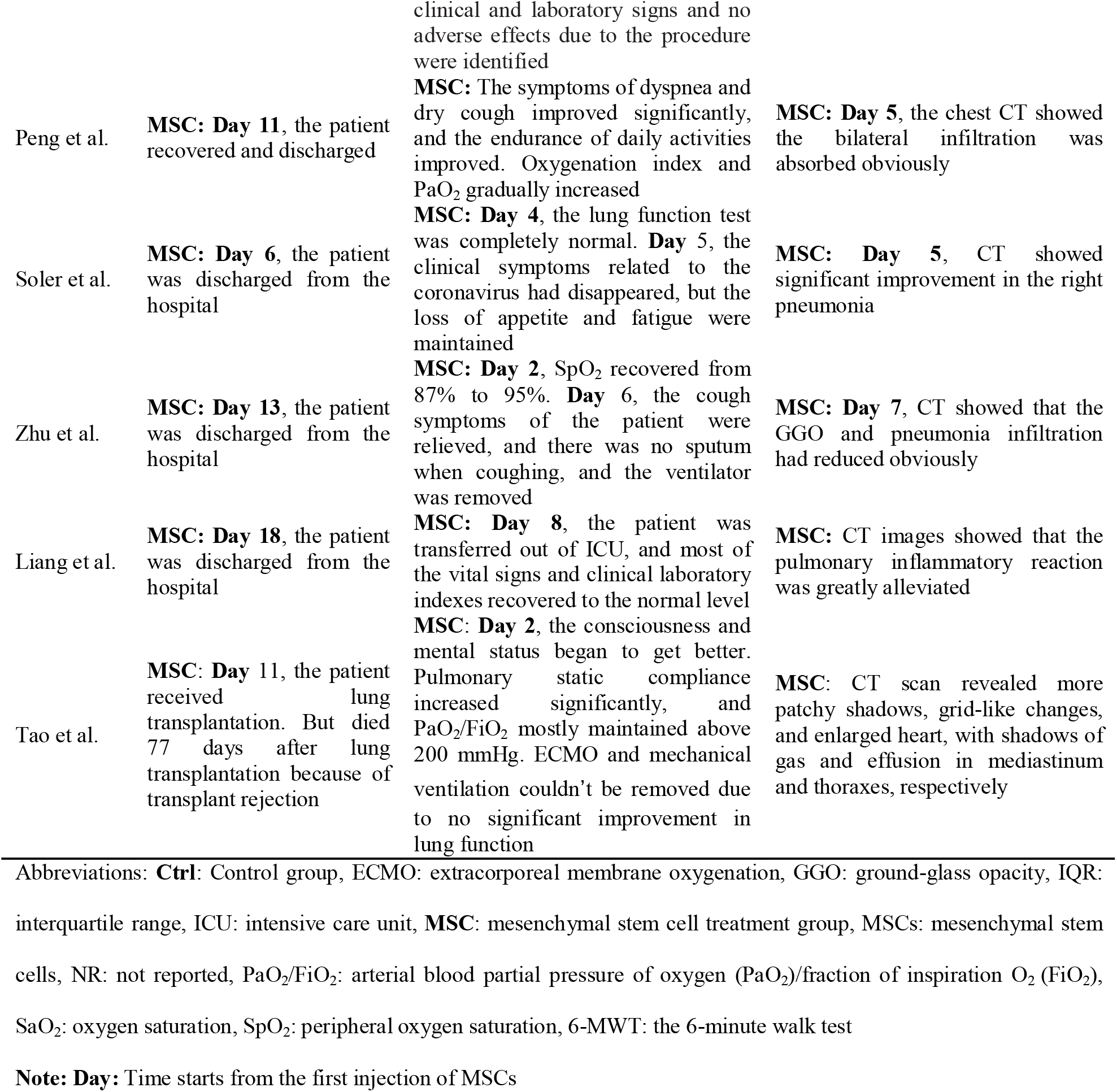
Clinical symptoms and imaging outcomes

Changes in general clinical symptoms and pulmonary function (including oxygenation index and chest radiology examinations) after treatment were recorded in 21 studies (**Table 5**) [10,12,13,22-25,27-40]. Different studies had different assessment indicators for lung function, which made it more difficult to extract the outcomes. The general clinical symptoms and pulmonary function (such as SaO_2,_ PaO_2_/FiO_2,_ the oxygenation index, and chest CT or X-ray) were found to improve in the early days after MSCs treatment in most of the studies. Some studies reported that the clinical improvement of MSCs group showed a significant improvement than control group, and the oxygenation index of the MSCs group recovered to the normal range faster than control group [23,25,27], and CT scores, the number of lobes involved, ground-glass opacity, and consolidation, which reflected reduced lung inflammation of MSCs group, were significantly better than those of control group [22-25]. However, there was a study reported that the cough of MSCs group showed a significant improvement compared with control group at Day 1 after MSCs treatment but no difference was found at other time points; the expiratory dyspnea showed a significant improvement compared with control group at Day 1, 3, and 5, but no difference was found at Day 7, 14, and 30 [25]. There was also a study reported that the saturation of pulse oxygen significantly improved in survivors compared to non-survivors even if all patients received MSCs treatment [10].

#### 3.4.3 Laboratory outcomes

The laboratory outcomes were shown in **Table 6**. The negative status of HCoV-19 nucleic acid was evaluated in 11 studies [12,13,22,24,26,29,35-39]. The average time from receiving the first injection to the nucleic acid turned to be negative ranged from about 4 to 15.8 days for MSCs-treated patients. Comparisons between MSCs group and control group were made in two studies, and there was no significant difference between the two groups [24,26].

**Table 6.**
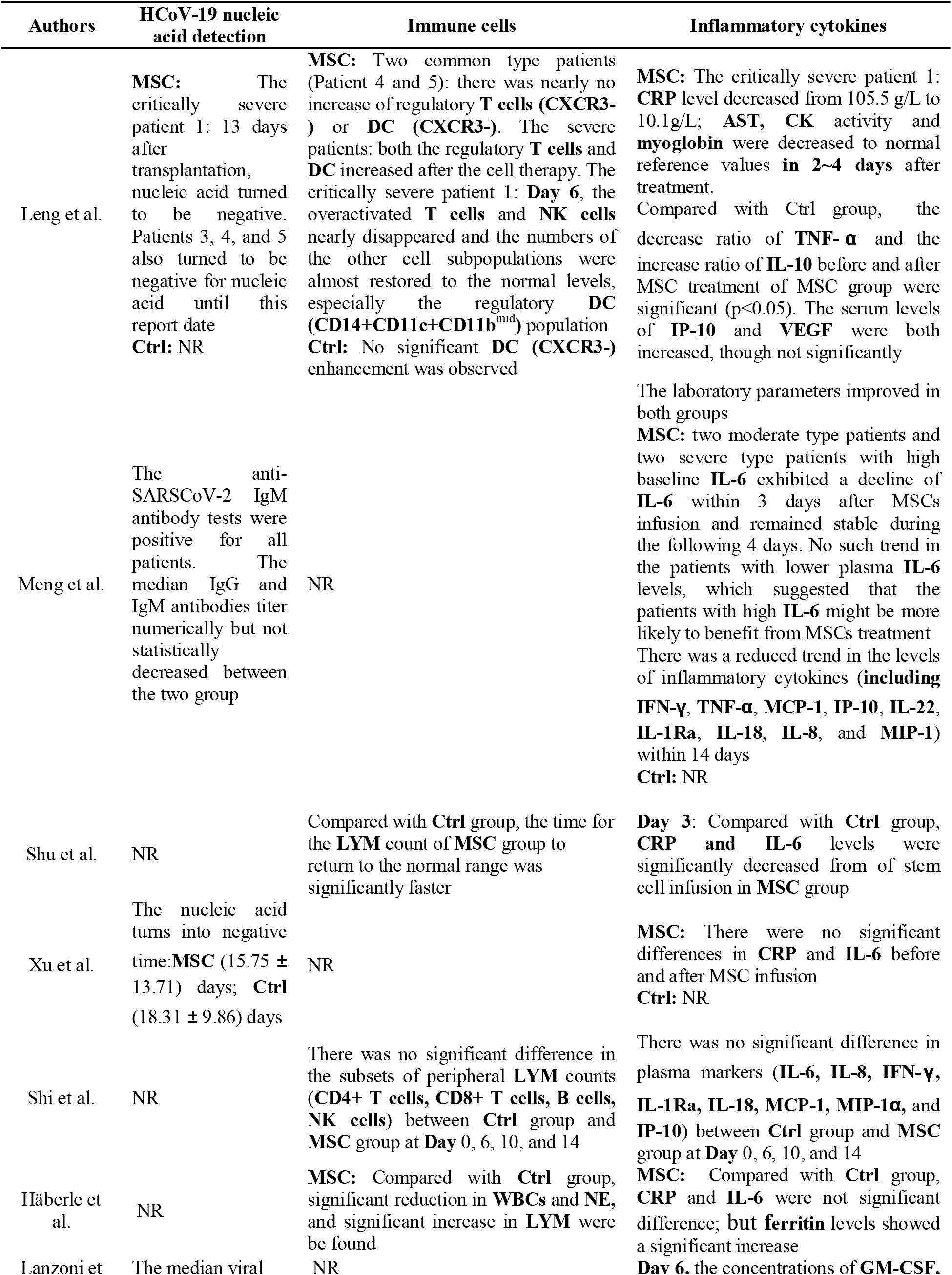
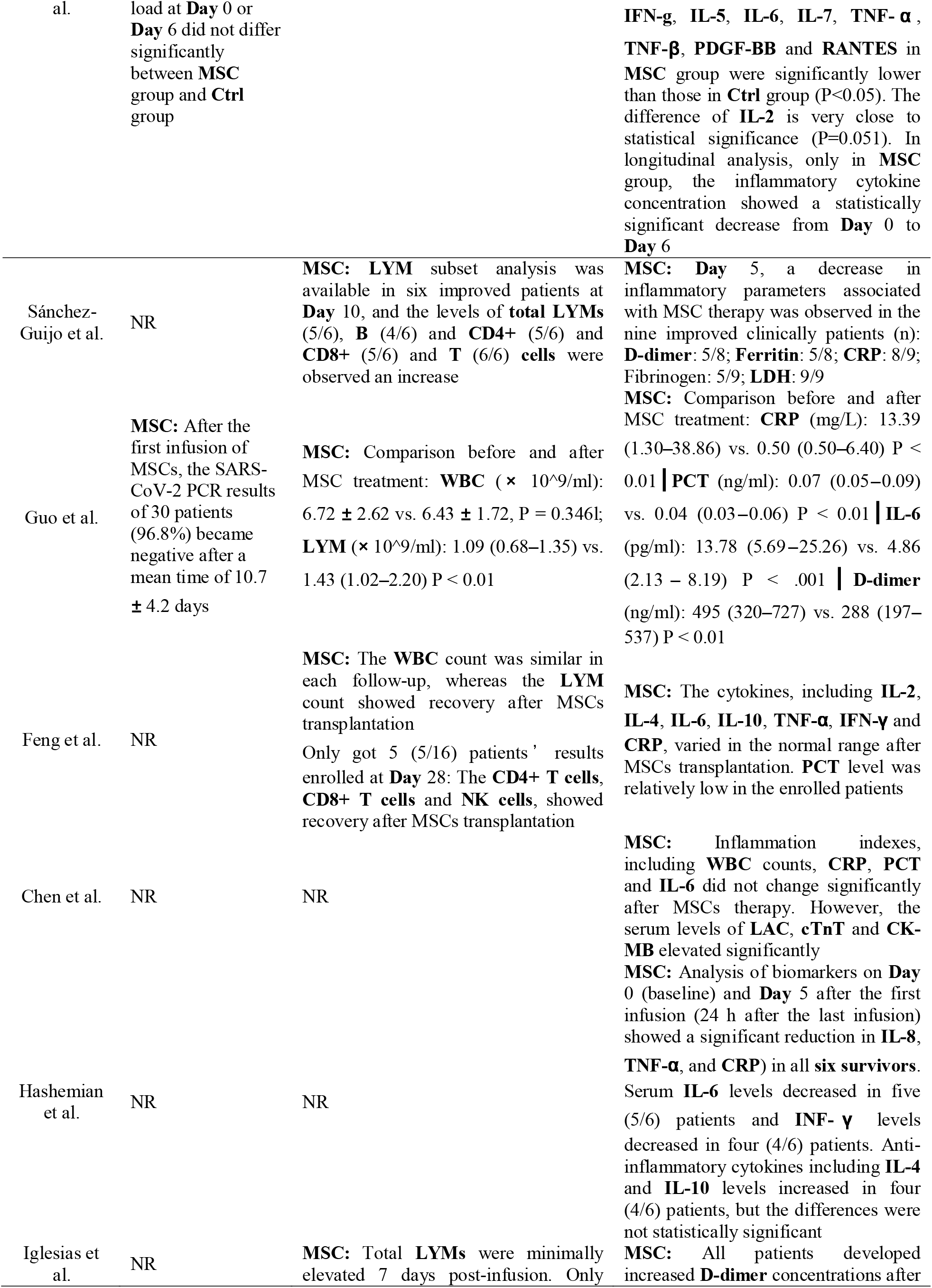
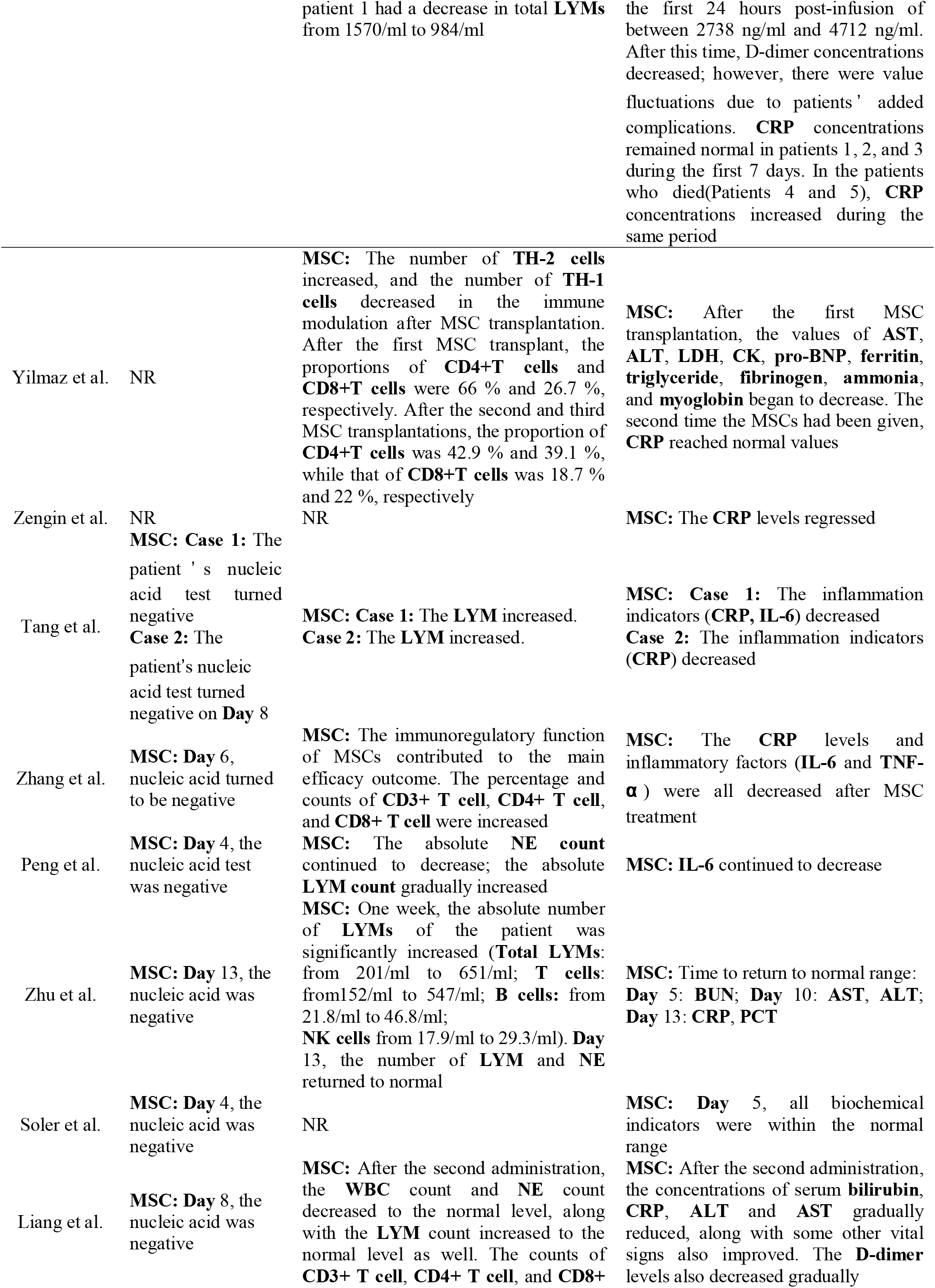
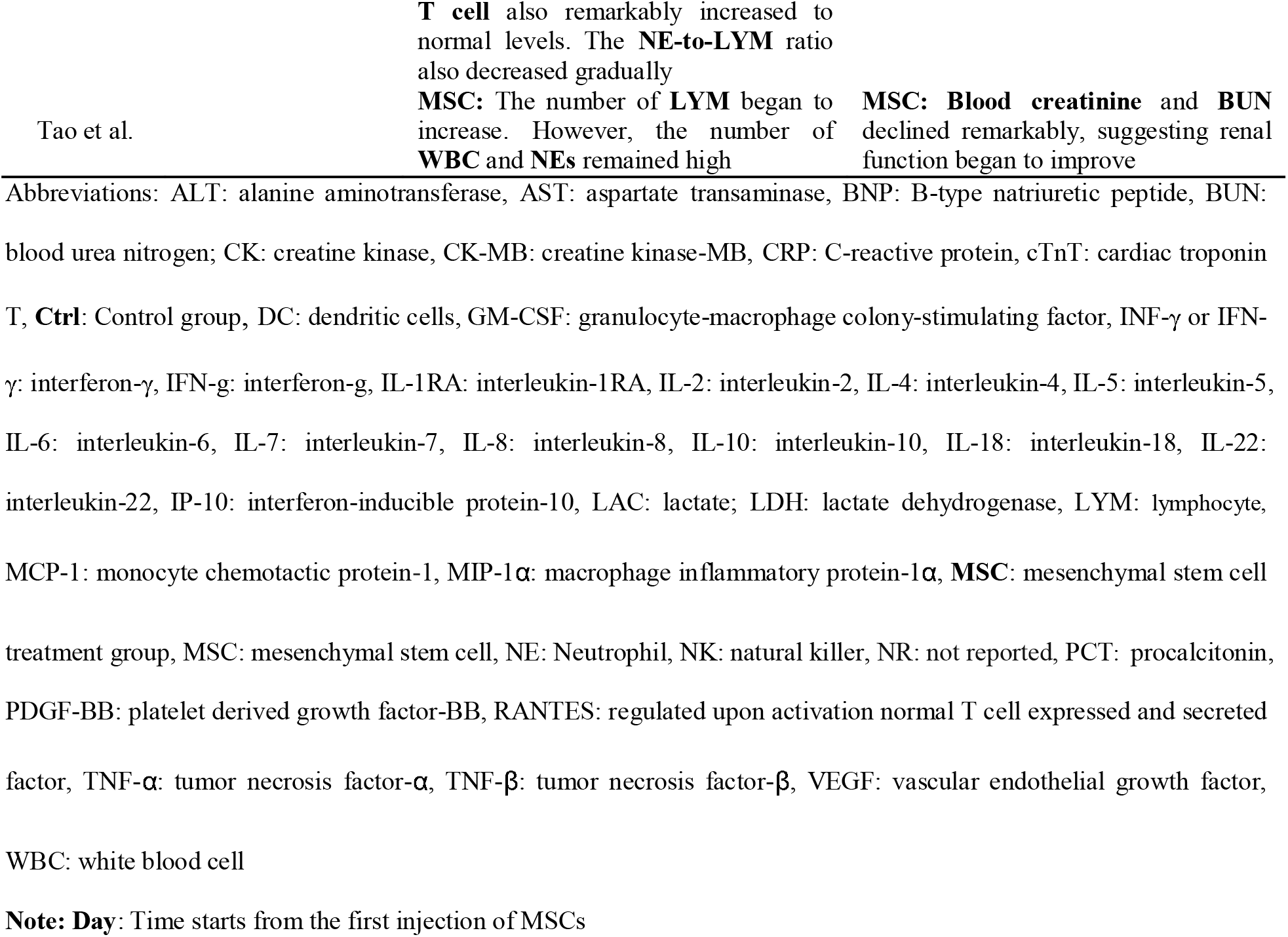
Laboratory outcome

The immune cells were tested in 15 studies [12,13,23,25,27-30,32,33,35,36,38-40]. The increased of DC, LYM, NK cells, T cells and B cells, and the decrease of NE and WBC were found in most of the studies. While Guo et al reported that there was no significant difference in WBC before and after MSCs treatment [29], and Iglesias et al reported that only one patient (1/5) had a decrease in total LYM from 1570/ml to 984/ml at 7 days post-infusion[32]. In controlled studies, a significant reduction in WBC and NE, and a significant increase in LYM were be found in MSCs group compared with control group [23,27]. However, there was a study reported that there was no significant difference in the subsets of peripheral LYM counts (CD4+ T cells, CD8+ T cells, B cells, NK cells) between the two groups at Day 0, 6, 10, and 14 after the first MSCs injection [25].

The inflammatory cytokines were evaluated in all studies. Most of the studies found that serum cytokines, chemokines, and growth factors improve to varying degrees following MSCs therapy. Two controlled studies reported that the concentrations of CRP, GM-CSF, IFN-g, IL-5, IL-6, IL-7, TNF-α, TNF-β, PDGF-BB and RANTES in MSCs group were significantly lower than those in control group [23,26]. However, we also noticed that there was no significant difference in plasma markers (IL-6, IL-8, IFN-γ, IL-1Ra, IL-18, MCP-1, MIP-1α, and IP-10) between the two groups reported in two controlled studies [25,27].

## 4. Discussion

It is exciting that some specific vaccines against the SARS-CoV-2 virus have been produced and promoted to vaccinate. In mainland China, people are being vaccinated for free in a planned way, and a total of 1 095.902 million doses of SARS-CoV-2 vaccines have been vaccinated as of June 23, 2021 [41]. However, the shortage of vaccines vaccine-related AEs and virus variant strains have kept some countries and regions in the shadow of the raging COVID-19 epidemic [42-46]. On this basis, other active treatments are still essential. At present, a considerable number of studies have analyzed the possibility of MSCs in the treatment of COVID-19, and some clinical trial results have confirmed its effectiveness. Therefore, this meta-analysis inspected the published results of MSCs treatment of COVID-19 patients, with focused analysis on safety, efficacy, and related pulmonary responses and immunologic. Our analysis of the studies found that MSCs therapy is safe and shows the potential to extenuate the damage of immunity and inflammation to the lungs and other organs and decrease the mortality of COVID-19 patients.

### 4.1 Safety

Safety is the primary concern for any new therapies, especially in patients at high risk of death due to treatment, and was carefully assessed for MSCs-treated patients in the reviewed studies. This study analyzed the occurrence of AEs and found that MSCs group had fewer patients with AEs than control group in five controlled studies [13,22,24-26], and the difference was statistically significant. Fifteen transient AEs related to MSCs treatment were reported [10,22,26,31,32], but most of them resolved spontaneously in a short time. Regrettably, 32 SAEs occurred in the 219 MSCs-treated patients, but the authors of these studies believed that none of these SAEs were found to be related to MSCs therapy [10,12,13,22-30,32-34,36-39]. Additionally, the number of SAEs of MSCs group was also less than that of control group in the controlled studies [13,22-27]. The safety is consistent with the experience of other human clinical trials involving MSCs treatment [47,48].

The mortality of patients infected with COVID-19 was used as the primary outcome to analyze the potential efficacy of MSCs therapy in this study. All seven controlled studies reported that MSCs group is associated with reduced mortality [13,22-27], and the pooled estimates of mortality showed that the mortality rate of MSCs group is significantly lower than that of control group (3.68% vs. 21.77%). The difference suggests that MSCs treatment may be effective in reducing mortality of these patients.

The COVID-19 disease accompanied with notable hematological manifestations, thrombocytopenia, and coagulation abnormalities on presentation and associated with poor outcomes during the disease courses [49]. There was a study reported that despite low molecular weight heparin prophylaxis or full anticoagulant therapy, the incidence of deep vein thrombosis, mainly asymptomatic, in hospitalized COVID-19 patients was 14.5% [50]. However, some patients had to suspend anticoagulation therapy due to bleeding or anemia and eventually died of hemodynamic disorders [32]. It is worth mentioning that 10 patients were already in very serious condition before receiving MSCs treatment and died of multiple organ failure or hemodynamic disorders [10,27,30,32,37]. Although their condition improved within a few days after MSCs transplantation, this sympathetic treatment failed to save their lives. Therefore, the authors of present study considered that although MSCs treatment could play a positive role in most COVID-19 patients and help save their lives, there are still individual differences in its efficacy.

### 4.2 Efficacy

The results of three studies showed that the average time of improvement (or recovery) in MSCs group was significantly shorter than that of control group [23,24,26], while two studies showed that the length of hospital stay was not different between the two groups [22,24]. The authors of present study believed that the reason for this difference was that the time from the onset of symptoms to the diagnosis of SARS-CoV-2 or hospitalization of patients was different in these studies. Therefore, it is not believed that the difference in the length of hospital stay is statistically significant in these studies with a small number of included cases and no uniform admission criteria.

Hashemian et al reported that non-survivors did not get the same amelioration in saturation of pulse oxygen as survivors even though all patients received MSCs treatment [10]. A case report showed that the patient’s consciousness and mental state began to improve after MSCs treatment, and his pulmonary compliance increased significantly, but extracorporeal membrane oxygenation (ECMO) and mechanical ventilation could not be resolved due to no significant improvement in lung function and chest CT scans. The treatment results were not satisfactory even after receiving five times MSCs injections, the patient was lucky enough to receive lung transplantation but unfortunately died of transplant rejection [40]. However, the general clinical symptoms, pulmonary function and radiographic imaging were found to ameliorate at the early days after MSCs treatment in most of the included studies, and the improvement in MSCs group was significantly better than that of control group. It has been found that intravenously infused MSCs migrate directly to the lung, where they can secrete a variety of factors, which play an important role in immune regulation, protection of alveolar epithelial cells, resistance to pulmonary fibrosis, and amelioration of lung function. It shows great benefits for the treatment of severe lung diseases in COVID-19 [51,52]. The findings of this study also further support the consideration of using MSCs to treat COVID-19-related pulmonary function decline.

The occurrence and development of SARS-CoV-2 depend on the interaction between virus infection and the immune system. Dysregulation of immune system in COVID-19 patients can contribute to serious illness. Dysregulation of the immune system such as lymphopenia and cytokine storm could be a crucial factor related to the severity of COVID-19 [49,53]. Compared with moderate COVID-19 cases, severe cases more frequently had dyspnea, lymphopenia, and hypoalbuminemia, with higher levels of ALT, LDH, CRP, ferritin, and D-dimer as well as markedly higher levels of IL-2R, IL-6, IL-10, and TNF-α. Absolute numbers of T cells, CD4^+^ T cells, and CD8^+^ T cells decreased in nearly all the patients and were markedly lower in severe cases than moderate cases [20]. In severe cases, patients suffer from ARDS, which is usually associated with elevated levels of inflammatory cytokines. Therefore, rebalancing the high inflammatory response of the host immune system and the regeneration of damaged cells seems to be the main way to treat this disease [54]. The main mechanism of MSCs therapeutic effect is due to the secretion of soluble factors, such as cytokines, chemokines, angiogenic factors, growth factors, and exosomes and extracellular vesicles. It is these complex mechanisms making MSCs suitable for treating complex and multifactorial diseases for which no other reductionistic drug treatments are available yet, such as COVID-19-related ARDS and other similar inflammatory diseases that involve a cytokine storm [14,55,56]. The present study found that MSCs therapy has a positive impact on the immune and inflammatory processes that lead to organ damage in COVID-19 patients. Most of the included studies showed that the number of immune cells ameliorated and serum cytoinflammatory factors gradually abated with MSCs treatment. Additionally, the WBC of patients had been corrected to the normal range before receiving MSCs injection, so there was no statistical difference before and after MSCs treatment in Guo’s study [29].

### 4.3 Other findings

ACE2 is the main host cell receptor for SARS-CoV-2 entry, and the virus uses the host cell transmembrane serine protease II (TMPRSS2) for Spike envelope protein priming [57]. It is known that ACE2 and TMPRSS2 are present on the surface of a variety of human cells, such as alveolar cells and capillary endothelium, while immune cells, such as T cells and B cells, and macrophage are negative for ACE2 [58]. There were studies reported that human MSCs do express neither ACE2 nor TMPRSS2, and human MSCs derived from fetal and adult tissues are not permissive to SARS-CoV-2 infection [13,59].

We also find out about comorbidities that the three most common comorbidities are hypertension, diabetes mellitus and obesity, which are the same as the concerns of many previous authors [60-63]. As the course of diabetes mellitus prolongs and some elderly hypertension patients need to be treated with ACE inhibitors which may increase the expression of ACE2. In addition, it has been found that pioglitazone and liraglutide, which are used to control blood glucose level and up-regulate the expression of ACE2 in experimental models [64-66]. Previous studies also confirmed that comorbidities such as hypertension, diabetes, and obesity are risk factors for severe symptoms and increased mortality in COVID-19 patients [60-64,67].

### 4.4 Research significance

This comprehensive systematic review and meta-analysis of published reports of MSCs therapy for COVID-19 have yielded several important findings. Primarily, the number of patients with AEs in MSCs group was significantly less than that in control group, and the mortality rate was also significantly slashed (3.68% vs. 21.77%). Only 36 SAEs occurred in 219 patients treated with MSCs, and none of them was related to MSCs transplantation. Additionally, MSCs therapy plays an active role in restoring lung function and improving symptoms. Furthermore, MSCs therapy is beneficial in coping with cytokine storms and correcting immune disorders. This study provides a sympathetic treatment option for doctors and patients in areas still under the shadow of the COVID-19 outbreak and accumulates experience for coping with new challenges in the future.

### 4.5 Limitations

Towards the end of this project, there are still not many clinical reports on MSCs for the treatment of COVID-19. Therefore, the limitations of this systematic review include the lack of large-scale RCTs, and no canonical treatment program and evaluation standard for MSCs. There are differences in the source, dose, activity, frequency, and inoculation interval of the MSCs in the included studies. It is unknown whether these will affect the treatment effect. Moreover, there may exist selection bias and insufficient description of the evaluation in published results.

## 5 Conclusion

This systematic review and meta-analysis of existing studies demonstrated the safety and effectiveness of MSCs in the treatment of COVID-19. There is an urgent need for adequately powered clinical trials to test the clinical results of MSCs therapy on patients with COVID-19 syndrome and SARS-CoV-2 infection, and to explore a standard MSCs therapy program.

## Data Availability

All data generated or analyzed during this study are included in this published article and its supplementary information files.

## Declarations

### Authors’ contributions

**Wang Junwu**: Material preparation, Data collection and analysis, Methodology, Formal analysis, investigation, and Writing - original draft. **Shi Pengzhi**: Conceptualization, Material preparation, Data collection and analysis, Methodology, Formal analysis and investigation. **Chen Dong**: Material preparation, Data collection and analysis, Methodology, Formal analysis and investigation. **Wang Shuguang**: Material preparation, Data collection and analysis, Methodology, Formal analysis and investigation. **Wang Pingchuan**: Material preparation, Data collection and analysis, Formal analysis and investigation. **Feng Xinmin**: Conceptualization, Methodology, Writing - review and editing, Supervision, Resources. **Zhang Liang**: Conceptualization, Methodology, Writing - review and editing, Funding acquisition, Resources, Supervision. All authors contributed to the study conception and design.

### Ethics approval and consent to participate

Not applicable.

