## Supplementary material for "The safety and efficacy of mesenchymal stem cells in the treatment of COVID-19-associated pneumonia: a systematic review and meta-analysis"

**Table S1** Study quality assessment tools

| Criteria | Yes/No/Other(CD, NR, NA) |
| --- | --- |
| **(1) Quality Assessment of Controlled Intervention Studies** |  |
| 1. Was the study described as randomized, a randomized trial, a randomized clinical trial, or an RCT? |  |
| 2. Was the method of randomization adequate (i.e., use of randomly generated assignment)? |  |
| 3. Was the treatment allocation concealed (so that assignments could not be predicted)? |  |
| 4. Were study participants and providers blinded to treatment group assignment? |  |
| 5. Were the people assessing the outcomes blinded to the participants' group assignments? |  |
| 6. Were the groups similar at baseline on important characteristics that could affect outcomes (e.g., demographics, risk factors, co-morbid conditions)? |  |
| 7. Was the overall drop-out rate from the study at endpoint 20% or lower of the number allocated to treatment? |  |
| 8. Was the differential drop-out rate (between treatment groups) at endpoint 15 percentage points or lower? |  |
| 9. Was there high adherence to the intervention protocols for each treatment group? |  |
| 10. Were other interventions avoided or similar in the groups (e.g., similar background treatments)? |  |
| 11. Were outcomes assessed using valid and reliable measures, implemented consistently across all study participants? |  |
| 12. Did the authors report that the sample size was sufficiently large to be able to detect a difference in the main outcome between groups with at least 80% power? |  |
| 13. Were outcomes reported or subgroups analyzed prespecified (i.e., identified before analyses were conducted)? |  |
| 14. Were all randomized participants analyzed in the group to which they were originally assigned, i.e., did they use an intention-to-treat analysis? |  |
| **(2)** **Quality Assessment of Case-Control Studies** |  |
| 1. Was the research question or objective in this paper clearly stated and appropriate? |  |
| 2. Was the study population clearly specified and defined? |  |
| 3. Did the authors include a sample size justification? |  |
| 4. Were controls selected or recruited from the same or similar population that gave rise to the cases (including the same timeframe)? |  |
| 5. Were the definitions, inclusion and exclusion criteria, algorithms or processes used to identify or select cases and controls valid, reliable, and implemented consistently across all study participants? |  |
| 6. Were the cases clearly defined and differentiated from controls? |  |
| 7. If less than 100 percent of eligible cases and/or controls were selected for the study, were the cases and/or controls randomly selected from those eligible? |  |
| 8. Was there use of concurrent controls? |  |
| 9. Were the investigators able to confirm that the exposure/risk occurred prior to the development of the condition or event that defined a participant as a case? |  |
| 10. Were the measures of exposure/risk clearly defined, valid, reliable, and implemented consistently (including the same time period) across all study participants? |  |
| 11. Were the assessors of exposure/risk blinded to the case or control status of participants? |  |
| 12. Were key potential confounding variables measured and adjusted statistically in the analyses? If matching was used, did the investigators account for matching during study analysis? |  |
| **(3) Quality Assessment Tool for Case Series Studies** | |
| 1. Was the study question or objective clearly stated? |  |
| 2. Was the study population clearly and fully described, including a case definition? |  |
| 3. Were the cases consecutive? |  |
| 4. Were the subjects comparable? |  |
| 5. Was the intervention clearly described? |  |
| 6. Were the outcome measures clearly defined, valid, reliable, and implemented consistently across all study participants? |  |
| 7. Was the length of follow-up adequate? |  |
| 8. Were the statistical methods well-described? |  |
| 9. Were the results well-described? |  |
| Quality Rating (Good, Fair, or Poor) |  |
| Rater #1 initials: |  |
| Rater #2 initials: |  |
| Additional Comments (If POOR, please state why): |  |
| CD, cannot determine; NA, not applicable; NR, not reported |  |

**Table S2** Quality assessment of studies

| **Authors** | **Study design** | **Q1** | **Q2** | **Q3** | **Q4** | **Q5** | **Q6** | **Q7** | **Q8** | **Q9** | **Q**  **10** | **Q**  **11** | **Q**  **12** | **Q**  **13** | **Q**  **14** | **Rater #1** | **Rater #2** | **Quality Rating** |
| --- | --- | --- | --- | --- | --- | --- | --- | --- | --- | --- | --- | --- | --- | --- | --- | --- | --- | --- |
| Leng et al. | CCT | Y | Y | NR | Y | Y | CD | CD | NR | Y | Y | NR | NR | - | - | F | F | F |
| Meng et al. | CCT | Y | Y | NR | Y | Y | Y | CD | NR | Y | Y | NR | Y | - | - | G | G | G |
| Shu et al. | RCT | Y | CD | CD | CD | CD | Y | Y | Y | Y | Y | Y | NR | Y | Y | G | G | G |
| Xu et al. | CCT | Y | Y | NR | Y | Y | Y | CD | NR | Y | Y | NR | Y | - | - | G | G | G |
| Shi et al. | RCT | Y | Y | Y | Y | Y | Y | Y | Y | Y | Y | Y | NR | Y | Y | G | G | G |
| Lanzoni et al. | RCT | Y | Y | Y | Y | Y | Y | Y | Y | Y | Y | Y | NR | Y | Y | G | G | G |
| Häberle et al. | CCT | Y | Y | NR | Y | Y | NR | CD | NR | Y | CD | NR | NR | - | - | F | F | F |
| Sánchez-Guijo et al. | Case series | Y | Y | Y | NA | CD | Y | Y | CD | CD | - | - | - | - | - | F | F | F |
| Guo et al. | Research Letter | Y | Y | Y | NA | CD | Y | CD | Y | CD | - | - | - | - | - | F | F | F |
| Feng et al. | Case series | Y | Y | Y | NA | CD | Y | Y | CD | CD | - | - | - | - | - | F | F | F |
| Chen et al. | Letter | Y | Y | Y | NA | CD | Y | CD | CD | Y | - | - | - | - | - | F | F | F |
| Hashemian et al. | Case series | Y | Y | Y | NA | Y | Y | Y | CD | Y | - | - | - | - | - | G | G | G |
| Iglesias et al. | Case series | Y | Y | Y | NA | Y | Y | CD | CD | Y | - | - | - | - | - | G | G | G |
| Yilmaz et al. | Case report | Y | Y | NA | NA | CD | Y | Y | CD | CD | - | - | - | - | - | F | F | F |
| Zengin et al. | Case report | Y | Y | NA | NA | CD | CD | Y | CD | CD | - | - | - | - | - | F | F | F |
| Tang et al. | Case report | Y | Y | NA | NA | Y | Y | Y | CD | Y | - | - | - | - | - | G | G | G |
| Zhang et al. | Case report | Y | Y | NA | NA | Y | Y | Y | CD | Y | - | - | - | - | - | G | G | G |
| Peng et al. | Case report | Y | Y | NA | NA | Y | Y | Y | CD | Y | - | - | - | - | - | G | G | G |
| Soler et al. | Letter | Y | Y | NA | NA | CD | CD | Y | CD | CD | - | - | - | - | - | F | F | F |
| Zhu et al. | Case report | Y | Y | NA | NA | CD | Y | Y | CD | Y | - | - | - | - | - | G | G | G |
| Liang et al. | Case report | Y | Y | NA | NA | Y | Y | Y | CD | Y | - | - | - | - | - | G | G | G |
| Tao et al. | Case report | Y | Y | NA | NA | CD | Y | Y | CD | Y | - | - | - | - | - | G | F | F |

Abbreviations: Q, question; Y, yes; CD, cannot determine; NA, not applicable; NR, not reported, G, good; F, fair
